# New insights into the genetic etiology of 57 essential and non-essential trace elements in humans

**DOI:** 10.1101/2023.04.25.23289097

**Authors:** Marta R. Moksnes, Ailin F. Hansen, Brooke N. Wolford, Laurent F. Thomas, Humaira Rasheed, Anica Simić, Laxmi Bhatta, Anne Lise Brantsæter, Ida Surakka, Wei Zhou, Per Magnus, Pål R. Njølstad, Ole A. Andreassen, Tore Syversen, Jie Zheng, Lars G. Fritsche, David M. Evans, Nicole M. Warrington, Therese H. Nøst, Bjørn Olav Åsvold, Trond Peder Flaten, Cristen J. Willer, Kristian Hveem, Ben M. Brumpton

## Abstract

Trace elements are important for human health but may exert toxic or adverse effects. Mechanisms of uptake, distribution, metabolism, and excretion are partly under genetic control but have not yet been extensively mapped. Here we report a comprehensive multi-element genome-wide association study (GWAS) of 57 essential and non-essential trace elements. We performed GWA meta-analyses of 14 trace elements in up to 6580 Scandinavian whole-blood samples, and GWASs of 43 trace elements in up to 2819 samples measured only in the Trøndelag Health Study (HUNT). We identified 11 novel genetic loci associated with blood concentrations of arsenic, cadmium, manganese, selenium, and zinc in genome-wide meta-analyses. In HUNT, several genome-wide significant loci were also indicated for other trace elements. Using two-sample Mendelian randomization, we found several indications of weak to moderate effects on health outcomes, the most precise being a weak harmful effect of increased zinc on prostate cancer. However, independent validation is needed. Our new understanding of trace element-associated genetic variants may help establish consequences of trace elements on human health.

## Introduction

Trace elements are present in living organisms at low concentrations, often defined as concentrations below 100 parts per million (ppm) or less than 100 μg/g^1^. Essential trace elements are vital for growth, development and normal physiology and biochemistry, and external sources are necessary as the body is unable to synthesize them. Other trace elements are classified as non-essential or toxic^2^. The major sources of exposure to trace elements in the general population are food, water, and air. Trace elements therefore often show considerable geographical variations^3^. Essential trace elements may also be toxic at elevated concentrations, and their uptake and metabolism are generally tightly regulated in the body^4–6^.

The mechanisms underlying variation in trace element concentrations between individuals are far from fully understood. Variations in trace element concentrations have previously been considered to be governed mainly by dietary intakes of food items with elevated concentrations^7^ and environmental exposure (e.g. geochemical variations, work-related exposure and anthropogenic pollution)^8, 9^, but genetic variation may also be an important factor^10–16^. A well-known example is body iron, where protein-altering mutations in the *HFE* gene cause a severe iron overload condition, hemochromatosis^17^. Knowledge about genetic factors controlling trace element concentrations may shed light on the biological pathways of trace elements in the body.

Few genome-wide association studies (GWAS) have investigated trace element concentrations and no previous studies have examined a comprehensive multielement panel of trace elements. Early twin studies have shown evidence for heritable variation in humans for copper, selenium, zinc, arsenic, cadmium, lead and mercury^10, 18^ and recent GWASs have reported genetic associations with blood concentrations of manganese, copper, selenium, zinc, lead, cadmium and mercury^12–16, 19^. However, many of the previous studies were limited by sample size or investigated only one or a few trace elements. Among the trace elements we investigate here, no published GWAS was found for 39 of them as of today.

Essential as well as non-essential trace elements have been linked to several health-related outcomes in humans, including neurodegenerative disorders^20–32^, autoimmune diseases^33–35^, endocrinological diseases^36–40^, cancers^41–43^ and bone health^44–47^. However, there is inconsistent and sometimes conflicting evidence for a protective or harmful effect of the trace elements on different diseases. Most of these studies have been observational and so the findings may have been influenced by confounding factors. Mendelian randomization (MR) methods have been developed to obtain an additional level of evidence for causal relationships. The potential causality of increased trace element levels on various diseases may be explored using genetic instruments for the individual elements. Trace elements have been measured in whole blood in two large Norwegian population-based studies: the Trøndelag Health Study (HUNT)^8, 48–51^, and the Norwegian Mother, Father, and Child Cohort Study (MoBa)^7, 52–54^. Further, genetic variants have been genotyped genome-wide and imputed using the Haplotype Reference Consortium reference panel (v1.1) in both cohorts^55^. GWASs of trace elements have also been reported in other cohorts, including the Swedish Prospective Investigation of the Vasculature in Uppsala Seniors (PIVUS) study^15^. The main aim of the current study was to identify genetic variants associated with whole blood trace element concentrations, and to explore potential causal associations between trace elements and cancers, neurodegenerative, autoimmune, endocrinological and bone related health outcomes. We therefore performed GWASs in HUNT and MoBa followed by a meta-analysis with the PIVUS study^15^. Further, we used the genetic associations for causal inference with Mendelian randomization, thereby estimating effects of circulating trace elements on health-related outcomes highlighted in previous literature.

## Results

### Genetic loci associated with trace elements

We identified 20 independent genetic loci (among 21 associations) that reached the genome-wide significance threshold (p-value < 5×10^−8^) in the meta-analyses of blood-concentrations of 14 trace elements measured in the HUNT (14 trace elements, sample size N=2819), MoBa (11 trace elements, N=2812) and PIVUS (11 trace elements, N=949) studies (Supplementary Table 1). The loci were consistently associated (i.e. same direction of the effect) with concentrations of essential (copper [number of variants, n=1], manganese [n=10], selenium [n=2], and zinc [n=3]) and toxic (arsenic [n=1], cadmium [n=3], and lead [n=1]) trace elements across the meta-analyzed cohorts (Table 1, Supplementary Tables 2 and 3, Supplementary Figures 1-14). Ten of these associations had previously been reported^12–16, 56, 57^ (one locus was associated with both manganese and cadmium levels), and 11 had not. Additionally, in HUNT alone we analyzed 43 additional trace elements that we could not meta-analyze because they had not been measured or the associated genetic variants were not tested in any other cohort. Here, we identified 27 genetic loci for 12 trace elements in the analysis (Supplementary Table 2, Supplementary Figures 15-38). Among these, we observed a common variant in *MORC4* with strong evidence of association with strontium concentrations (minor allele frequency (MAF) =0.19, p-value=2.0×10^−16^), and a common variant in *HK1* that was associated with magnesium concentrations (MAF=0.14, p-value=2.2×10^−8^). One locus in *SLC18A2* was associated with selenium in HUNT (MAF=0.27, p-value=3.8×10^−8^), but the index (lowest p-value) variant was not tested in MoBa, and none of the tested proxy variants (in high linkage disequilibrium (LD) in HUNT, i.e. correlation r^2^_HUNT_>0.8 with the index variant) were associated with selenium concentrations in MoBa. Most of the other variants from HUNT only were low-frequency (MAF<5%) or rare (MAF<0.5%) and given that they were only tested for association in one cohort, they were more likely to be false positive findings. Sensitivity analyses correcting for fish intake (arsenic, selenium), smoking status (arsenic, cadmium, copper, lead) and weekly alcohol intake (iron, manganese, zinc, lead) in HUNT did not substantially change the effect sizes of the index variants (Supplementary Table 4).

**Table 1:** Index variants in genetic loci associated (p-value<5E-8) with trace element concentrations in meta-analyses

| Trace Element | Locus novelty | RsID | Chr | Pos (GRCh37) | Ref | Alt | AF | Effect | SE | P-value | N | Nearest gene | Consequence |
| --- | --- | --- | --- | --- | --- | --- | --- | --- | --- | --- | --- | --- | --- |
| As | Novel | rs73060324 | 3 | 45785915 | T | G | 0.10 | 0.25 | 0.03 | 4.8×10<sup>-16</sup> | 5591* | *SACM1L* | Nonsynonymous** |
| Cd | Known<sup>56</sup> | rs2054394 | 4 | 103268763 | G | A | 0.11 | -0.21 | 0.03 | 4.9×10<sup>-14</sup> | 6542 | *SLC39A8*,<br>*LOC105377621* |  |
| Cd | Known<sup>16</sup> | rs6987313 | 8 | 71570989 | T | C | 0.52 | -0.11 | 0.02 | 5.2×10<sup>-11</sup> | 6542 | *LACTB2-AS1* |  |
| Cd | Novel | rs953733 | 15 | 45393667 | A | G | 0.08 | 0.21 | 0.03 | 2.9×10<sup>-10</sup> | 6542 | *DUOX2* | Nonsynonymous*** |
| Cu | Known<sup>14,19</sup> | rs34004251 | 3 | 148929951 | T | A | 0.20 | 0.21 | 0.02 | 4.7×10<sup>-23</sup> | 6564 | *CP* |  |
| Pb | Known<sup>13</sup> | rs1805312 | 9 | 116152990 | C | G | 0.08 | -0.30 | 0.03 | 7.1×10<sup>-19</sup> | 6564 | *ALAD* |  |
| Mn | Known<sup>15</sup> | rs6541114 | 1 | 220074279 | G | T | 0.19 | 0.47 | 0.02 | 6.3×10<sup>-99</sup> | 6564 | *ZC3H11B*;<br>*SLC30A10* |  |
| Mn | Known<sup>15</sup> | rs13107325 | 4 | 103188709 | C | T | 0.05 | -0.42 | 0.04 | 6.0×10<sup>-25</sup> | 6564 | *SLC39A8* | Nonsynonymous |
| Mn | Novel | rs467304 | 6 | 5119696 | C | T | 0.31 | 0.15 | 0.02 | 6.5×10<sup>-14</sup> | 6564 | *LYRM4-AS1* |  |
| Mn | Known<sup>57</sup> | rs1800562 | 6 | 26093141 | G | A | 0.08 | -0.22 | 0.03 | 4.5×10<sup>-12</sup> | 6564 | *HFE* | Nonsynonymous |
| Mn | Novel | rs4440696 | 9 | 88903367 | T | A | 0.65 | 0.15 | 0.02 | 2.6×10<sup>-16</sup> | 6564 | *TUT7* |  |
| Mn | Novel | rs7176565 | 15 | 65998702 | C | T | 0.76 | -0.16 | 0.02 | 5.5×10<sup>-13</sup> | 6564 | *DENND4A* |  |
| Mn | Novel | rs2272783 | 18 | 55238820 | A | G | 0.06 | 0.36 | 0.04 | 8.9×10<sup>-21</sup> | 6564 | *FECH* |  |
| Mn | Novel | rs6099115 | 20 | 54941140 | C | T | 0.18 | -0.16 | 0.02 | 1.1×10<sup>-12</sup> | 6564 | *FAM210B* | Nonsynonymous |
| Mn | Novel | rs5997397 | 22 | 29154455 | A | G | 0.44 | -0.11 | 0.02 | 1.2×10<sup>-9</sup> | 6564 | *HSCB* |  |
| Mn | Novel | rs855791 | 22 | 37462936 | A | G | 0.58 | -0.12 | 0.02 | $3.9 \times 10^{-12}$ | 6564 | *TMPRSS6* | Nonsynonymous |
| Se | Novel | rs1047891 | 2 | 211540507 | C | A | 0.31 | 0.11 | 0.02 | $6.0 \times 10^{-9}$ | 5615* | *CPS1* | Nonsynonymous |
| Se | Known<sup>12</sup> | rs17823744 | 5 | 78344976 | A | G | 0.12 | 0.25 | 0.03 | $3.3 \times 10^{-22}$ | 5615* | *DMGDH* | |
| Zn | Novel | rs322884 | 1 | 199029020 | A | C | 0.66 | -0.11 | 0.02 | $1.3 \times 10^{-9}$ | 6564 | *LINC01221* | |
| Zn | Known<sup>12</sup> | rs10097917 | 8 | 86260295 | A | T | 0.61 | -0.17 | 0.02 | $4.7 \times 10^{-22}$ | 6564 | *CA1* | |
| Zn | Known<sup>12</sup> | rs12591119 | 15 | 75355944 | A | G | 0.18 | -0.29 | 0.02 | $1.4 \times 10^{-38}$ | 6564 | *PPCDC; C15orf39* | |

### Protein-altering variants

Among the 20 top association signals, we identified eight protein-altering single nucleotide polymorphisms (SNPs) associated with trace elements: Four manganese associated index variants (rs13107325 in *SLC39A8,* rs1800562 in *HFE*, rs6099115 in *FAM210B* and rs855791 in *TMPRSS6*) and one selenium associated index variant (rs1047891 in *CPS1*) were nonsynonymous SNPs. Among these, only rs13107325 had previously been reported for manganese in a GWAS^15^. Although the hemochromatosis variant rs1800562 was not associated with manganese at the GWAS significance level in a previous GWAS^15^, women with *HFE* variants were reported to have 12% lower blood manganese concentrations in another study^57^. Further, three nonsynonymous SNPs were in strong LD (r^2^ > 0.8) with index variants: rs17279437 in *SLC6A20* (p-value=9.8×10^−16^, r^2^_HUNT_ = 0.88 with arsenic index variant rs7306032), rs269868 in *DUOX2* (p-value=5.7×10^−10^, r^2^_HUNT_=0.96 with cadmium index variant rs953733) and rs57659670 in *DUOX2* (p-value=3.2×10^−10^, r^2^_HUNT_=0.99 with cadmium index variant rs953733) (Supplementary Table 5).

### SNP heritability estimates and genetic and phenotypic correlations of trace elements

For the concentrations of 10 trace elements that had sample sizes above 5000 (cadmium, cobalt, copper, lead, manganese, mercury, molybdenum, selenium, thallium, and zinc), the estimated narrow-sense SNP heritability h^2^ ranged from h^2^=0.01±0.09 (thallium) to h^2^=0.29±0.10 (manganese). The estimates were higher for essential trace elements (h^2^ between 0.16 and 0.29) than for non-essential trace elements (h^2^ between 0.01 and 0.11) (Supplementary Table 6).

We estimated the genetic correlation between all pairs of these 10 trace elements and found absolute values of the genetic correlations to range from 0.01 (cobalt against lead and cobalt against mercury) to 0.97 (molybdenum against cadmium), although the estimates were imprecise and therefore mostly uninformative (Supplementary Figure 44, Supplementary Table 7). For comparison, we also estimated the phenotypic correlation (Supplementary Figure 39, Supplementary Table 7), where the absolute values ranged from around 0 (manganese against lead and manganese against mercury) to 0.45 (mercury against selenium). We did not observe patterns among the genetic or the phenotypic correlations related to the known ion binding preferences of the trace elements^58^, which we chose to group as class A (“oxygen-seeking”) (molybdenum, manganese, zinc), intermediate (cobalt, cadmium, copper), and class B (“sulfur/nitrogen-seeking”) (lead, mercury, selenium).

### Phenome-wide associations of trace element loci

We investigated the genetic relationship between trace element concentrations and other complex traits by examining associations of the index variants identified by the meta-analyses with 1326 phenotype codes (‘phecodes’^59^), 30 blood biomarkers and 167 other continuous traits and measurements of the UK Biobank. In total, 17 index variants from trace element meta-analyses were associated (p-value < 9.7×10^−7^, threshold Bonferroni corrected for 51 782 tests) with additional phenotypes (Figure 1, Supplementary Figure 40, Supplementary Tables 8-10):

**Figure 1:**
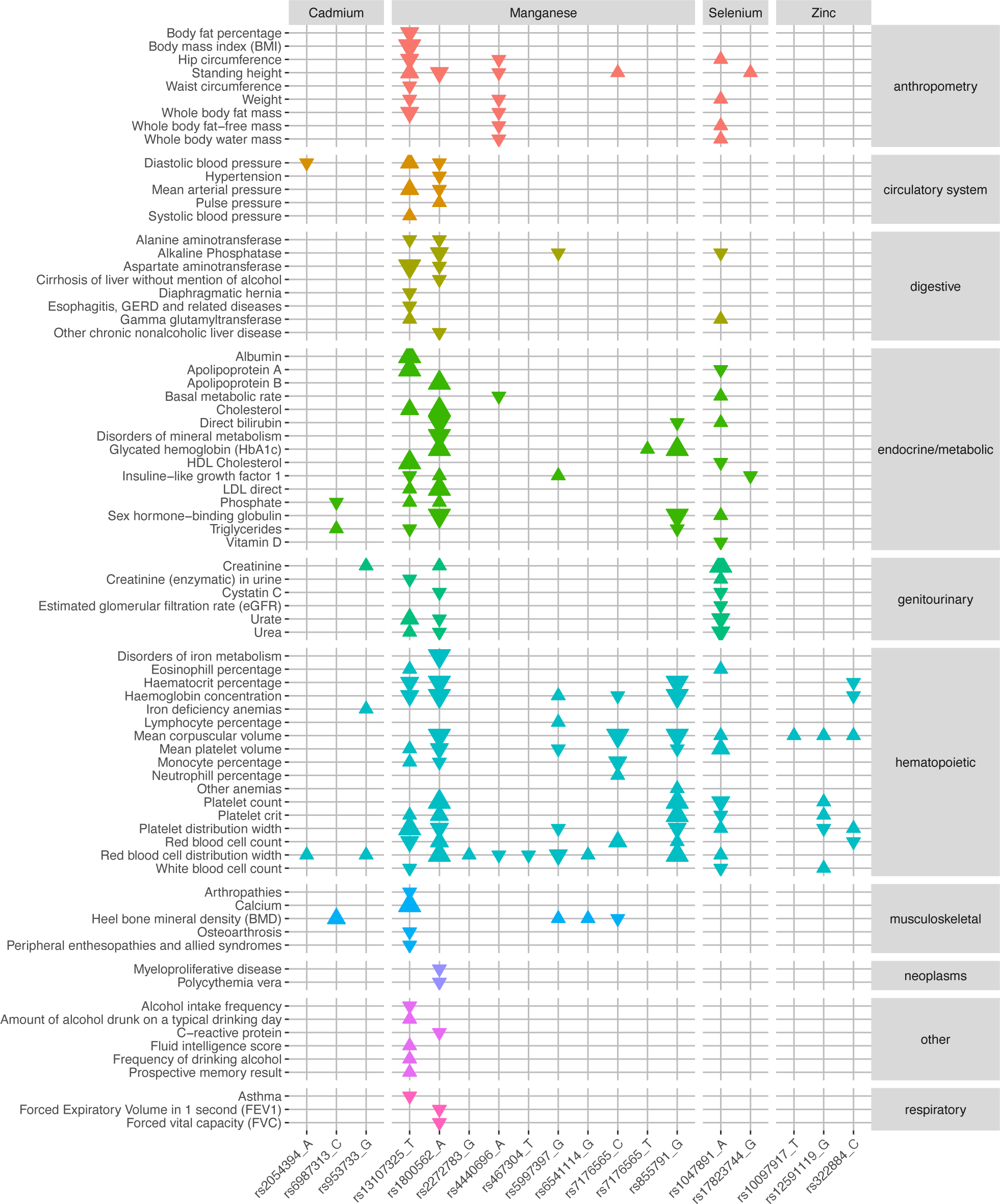
Phenome-wide associations between meta-analysis index variants and selected outcomes in the UK Biobank. Each triangle represents a statistically significant (p-value<9.7×10^−7^) association between an index variant from trace element GWA meta-analysis (x-axis, grouped by trace element) and an outcome in the UK Biobank (y-axis, grouped and colored according to biological domain). Larger triangles represent lower p-values, and the direction indicates if the direction of effect for the indicated allele is the same (up) or opposite (down) as the association with the trace element. Note: Figure 1 only presents a selection of the associated outcomes in UK Biobank. The full list of associations with meta-analysis index variants are visualized in Supplementary Figure 40.

Three nonsynonymous index variants for manganese (rs13107325 [*SLC39A8*], rs1800562 [*HFE*], rs855791 [*TMPRSS6*]) and one index variant for cadmium in high LD with nonsynonymous SNPs (rs953733 [*DUOX2*]) were associated with phecodes (Supplementary Table 8, Figure 1): The manganese decreasing variant in the metal ion transporter gene *SLC39A8* (rs13107325) was positively associated with diseases of the esophagus and with musculoskeletal conditions. The manganese decreasing, but iron increasing, hemochromatosis variant (rs1800562) was positively associated with disorders of mineral and iron metabolism, but also with several diseases in other biological domains. The manganese increasing and iron decreasing^60^ variant rs855791 was positively associated with other anemias. The cadmium increasing variant found in a locus that has also been associated with iron status biomarkers^61^ (rs73060324), was positively associated with iron deficiency anemias.

Further, eight index variants, representing three trace elements (cadmium, manganese, and selenium), were associated with blood biomarkers in the UK Biobank (Supplementary Table 9, Figure 1, Supplementary Figure 40): Five manganese variants (rs13107325, rs1800562, rs7176565, rs5997397 and rs855791) were associated primarily with endocrine/metabolic biomarkers, but also with digestive, genitourinary, musculoskeletal, and inflammatory biomarkers. The cadmium increasing variant rs953733 was positively associated with levels of creatinine, a marker for kidney function^62^, while the cadmium decreasing rs6987313 was associated with phosphate and triglyceride levels. The selenium increasing nonsynonymous SNP rs1047891 [*CPS1*]) was associated with endocrine/metabolic, genitourinary, and digestive biomarkers. Finally, the selenium increasing rs17823744 (*DMGDH*) was negatively associated with insulin-like growth factor 1 (IGF-1).

Among the GWAS results for continuous traits in the UK Biobank, we observed associations for 17 meta-analysis index variants (for cadmium, manganese, selenium, and zinc) (Supplementary Table 10, Figure 1, Supplementary Figure 40). The majority of the associations were with different blood cell indices, but cadmium, manganese and selenium variants were also associated with continuous traits from several other biological domains, including musculoskeletal and anthropometric measures (cadmium, manganese, selenium), blood pressure traits (cadmium and manganese), biomarkers in urine and variables derived from endocrine/metabolic blood biomarkers (manganese and selenium), measures of cognitive ability (manganese) and alcohol habits (manganese).

In addition to the associations with the meta-analysis index variants, we also assessed the phenome-wide associations with common and low-frequency (MAF>0.5%) variants identified only in HUNT. Here, four common variants (strontium [rs17326228], cesium [rs7785293], magnesium [rs16926246] and silicon [rs62228297]) were associated (p-value < 9.7×10^−7^) with blood biomarkers and/or continuous traits in the UK Biobank (Supplementary Tables 11-12, Supplementary Figure 41): The cesium and magnesium variants were associated with a range of blood cell related traits, the cesium and strontium variants with genitourinary biomarkers and measures, the magnesium variant with endocrine/metabolic biomarkers and measures, and the silicon variant with standing height.

### Mendelian Randomization

We used two-sample MR to perform an exploratory examination of potential causal effects of trace elements on health outcomes within neurodegenerative, autoimmune, and endocrinological diseases, cancers and bone related domains as highlighted in the literature^20–47^. Here, we used the index SNPs from the meta-analyses (manganese, selenium, zinc, copper, lead, arsenic, and cadmium) or from the GWASs in HUNT (strontium) as instruments for the selected trace elements (F-statistics>10 for all trace elements except cadmium [F-statistic=9]: Supplementary Table 13). For the SNP-outcome associations, we used summary-level data from large GWASs of selected neurodegenerative disorders (Alzheimer’s disease, Parkinson’s disease, and multiple sclerosis), autoimmune diseases (rheumatoid arthritis and autoimmune thyroid disorder), endocrinological diseases (hypothyroidism and type 2 diabetes), bone related traits and disorders (bone mineral density, bone fractures, osteoporosis) and cancers (prostate cancer and colorectal cancer). The sources of the summary-level data can be found in Supplementary Table 13. The most precise association indicated a weak causal effect of zinc on prostate cancer (odds ratio (OR)=1.06, 95% CI=1.01-1.12) (Supplemental Table 13). There were also stronger but less precise associations indicating a causal effect of arsenic and copper on Parkinson’s disease (OR=1.24, 95% CI=0.98-1.57 [arsenic], OR=1.17, 95% CI=0.89-1.54 [copper]), as well as protective effects of selenium on colorectal cancer (OR=0.89, 95% CI=0.75-1.07) and selenium, cadmium and lead on multiple sclerosis (OR=0.86,95% CI=0.64-1.15 [selenium], OR=0.86,95% CI=0.72-1.01 [cadmium], OR=0.70, 95% CI=0.52-0.95 [lead]) (Supplemental Table 13). The results should however be interpreted with caution due to limitations such as few SNPs per instrument and known correlations and common biological pathways of trace elements. Otherwise, the MR estimates were generally imprecise, and generally they did not give convincing evidence for causal effects of trace elements in the remaining outcomes (Supplementary Table 13).

## Discussion

In the present study, we investigated genetic variants associated with whole blood trace element concentrations, combining data from up to 6564 individuals in HUNT, MoBa and PIVUS. Our GWAS meta-analyses identified genetic contributions to whole blood concentrations of essential (copper, manganese, selenium, and zinc) and non-essential or toxic (arsenic, cadmium, and lead) trace elements. In total, 20 loci were associated with trace element concentrations across multiple cohorts, confirming 9 loci from previous studies and identifying 11 novel loci. Seven of the novel loci were associated with manganese concentrations, including two nonsynonymous index variants. Four novel genetic loci were associated with selenium, zinc, arsenic, and cadmium, among which we also identified nonsynonymous variants in high LD (r^2^>0.8) with the index variants. The genetic variants that we identified in the present study had small to moderate effect sizes. They could, however, still contribute to individual differences in trace element concentrations in combination with many other factors. Identification of these loci expands our knowledge about the genetic contribution to trace element concentrations and indicates proteins that could aid in establishing mechanisms for absorption, distribution, metabolism, and excretion.

Because essential trace elements are not produced in the body, but are necessary for normal physiology, we would expect genes associated with essential trace elements to encode proteins that include these elements or are involved in their respective regulatory processes. This is in line with our observations in the meta-analyses of copper, zinc, and manganese, where we replicated previously reported loci with index variants near or in genes encoding the copper-binding metalloprotein ceruloplasmin (*CP*)^63^, the metalloenzyme carbonic anhydrase 1 (*CA1*) involved in the zinc balance^64^, and two divalent metal ion transporters *SLC30A10*^65^ and *SLC39A8*^66^ (nonsynonymous variant), where deficiency in the latter is known to cause severe manganese deficiency^67^. Other nonsynonymous variants we observed had, to our knowledge, little or no known underlying biology related to the associated trace elements. For example, we observed a novel association between selenium and a nonsynonymous variant in the urea cycle gene *CPS1.* Although the variant is associated with vitamin D^68^, which has again been hypothesized to interact with selenium^69^, and the gene has been observed to be upregulated in mice fed excessive amounts of selenium^70^, the underlying biology of the association with selenium in humans is not established. Because some trace elements interact with each other, or have common uptake mechanisms and co-transport, an imbalance in the concentration of one trace element might also change the concentration of others^7, 50, 71, 72^: For example, manganese is partly transported by both zinc and iron transporters^73^, and low iron stores have been associated with higher blood concentrations of other trace elements^7, 50^. Further, functional variants in iron metabolism genes have been associated with lower blood manganese concentrations^57^, including rs1800562 in *HFE* that we replicated here. The novel loci we observed to be associated with manganese, as well as the *CPS1* locus for selenium, are in line with this pattern: One manganese associated nonsynonymous SNP is known in iron deficiency anemia (rs855791 in *TMPRSS6*)^74^. Further, the gene that is closest to the index variant in another locus is the interleukin-6 regulator gene *TUT7*^75^, which stimulates the iron regulator hepcidin^76^. The index variants in other novel loci are close to genes involved in mitochondrial iron uptake (*FAM210B*^77^, where we also find the nonsynonymous index variant rs6099115) or genes associated with hematological traits or involved in the synthesis of iron containing compounds (*LYRM4*, *FAM210B*, *DENND4A*, *FECH*, *HSCB*)^77–80^. The *CPS1* index variant (rs1047891) was in high LD (r^2^_HUNT_ =0.92) with an index variant (rs715) for total iron binding capacity in our previous study^61^. These loci were not associated with iron in the current study; however, the analysis of iron had limited statistical power with less than half the sample size of manganese.

Non-essential trace elements typically do not have their own transport proteins or specific mechanisms for metabolism and are generally taken up into the body using the routes of macronutrients^81^ or essential trace elements with similar chemical properties^71^. This is illustrated by one of the known loci we replicate for the toxic metal cadmium, where the index variant is nearest to the zinc/manganese transporter gene *SLC39A8*, which was also associated with manganese. For cadmium, we also report a novel locus with two nonsynonymous variants in *DUOX2*, both in high LD with the index variant. The gene codes for a thyroid hormone synthesis related protein^82^ with a heme binding site^83^ and was associated with iron status in previous GWASs^61, 84^. Similarly, the nonsynonymous variant rs17279437 in the glycine transporter gene *SLC6A20* has been associated with excessive glycine excretion^85^. In this study, the variant was in high LD with the index variant for arsenic, which could potentially indicate that the protein can also transport arsenic. Glycine levels have also been associated with arsenic exposure in mouse models^86^.

The genetic associations observed only in HUNT warrant further replication. One interesting result identified in HUNT was a selenium locus within a solute carrier gene (*SLC18A2*). Although the index variant was not tested in MoBa, variants in high LD (r^2^>0.8) with the index variant were tested, but not associated with selenium in that data. The non-association in MoBa could however potentially be a result of sex-specific or pregnancy-specific exposure patterns. Further, index variants in gold and magnesium loci were in or nearest to genes associated with hemoglobin and iron regulation (*CD163* and *HK1*)^87, 88^. The strong association between hypomagnesemia and type 2 diabetes^89^, combined with a previously reported association between the magnesium *HK1* index variant with levels of the type 2 diabetes biomarker glycated hemoglobin (HbA1c)^90^ could support this finding. However, associations between *HK1* and HbA1c could also reflect the erythrocyte lifetime regardless of diabetes status^91^. We also observed a strong association between strontium concentrations and a locus in *MORC4*. Strontium is chemically very similar to calcium. It has effects on bone balance^92^, and could potentially induce skeletal abnormalities in very high doses^92^. A rare genetic variant in *MORC4* has been associated with a 3.4 times increased risk of osteoarthritis (Open Targets Platform^93^, accessed 25.05.2022), and another *MORC* family member, *MORC3*, is involved in calcium homeostasis and maintenance of bone remodeling^94^. We therefore speculate that *MORC4* could have similar functions or be involved in similar pathways.

In the PheWAS analyses, we observed the well-known association of functional variants in iron metabolism genes (the hemochromatosis *HFE* variant, rs1800562, and the *TMPRSS6* variant, rs855791) with disorders of iron and mineral metabolism and anemias. As expected, and previously demonstrated in the UK Biobank^61^, rs1800562 was also associated with several known clinical manifestations of *HFE* hemochromatosis^95, 96^. Likewise, the index variant in the *DUOX2* (cadmium) locus known from GWASs of iron status biomarkers^61, 84, 97^, was associated with iron deficiency anemia. The many associations between trace element index variants and blood cell indices could also potentially reflect their correlation with iron status. The associations between digestive and musculoskeletal disorders and rs13107325 (manganese), which alters the metal ion transporter ZIP8 (encoded by *SLC39A8*), could potentially highlight the role of manganese or other divalent trace metals in these conditions. Associations with basal metabolic rate and metabolic biomarkers could possibly be related to the processing of trace elements in the body, and associations with biomarkers for liver and kidney function could be related to the clearance of trace elements that are toxic or in high abundance. However, population stratification could also have caused false associations with some of the variants.

We estimated the heritability of concentrations of 10 trace elements with meta-analysis sample size above 5000, ranging from low to moderately high heritability (0.01±0.09 [thallium] to 0.29±0.10 [manganese]). The lower heritability for non-essential trace elements compared to essential trace elements could reflect that humans have not evolved genes and biological pathways to handle non-essential trace elements. The very low heritability of thallium could also be because most of the samples from MoBa were below the detection limit. Further, many of the heritability estimates could be low because LD Score regression underestimates the heritability of traits that are not highly polygenic^98^.

The genetic and phenotypic correlations between nine trace elements did generally not correspond well to each other, however the genetic correlations were highly imprecise and therefore mostly uninformative. Mercury and selenium had the strongest phenotypic correlation, which is in line with the known mercury-selenium antagonistic relationship^99^. In general, the genetic correlations were higher than the phenotypic correlations, although the range of the different phenotypic correlation estimates were similar to those previously reported in MoBa^7^. This could potentially reflect a relatively low polygenicity of trace element concentrations, where few genetic loci might influence many different trace elements, while there are a variety of different factors influencing the phenotypic variations.

Using two-sample MR, we observed indications of a weak harmful effect of circulating zinc on prostate cancer. The effect of zinc on prostate cancer is debated, but a harmful effect has been found in some previous studies^100^. The results also gave weak evidence for a harmful effect of arsenic and copper in Parkinson’s disease, and for a protective effect of selenium on colorectal cancer and selenium, cadmium, and lead on multiple sclerosis. The apparently protective effect of lead on multiple sclerosis is not in line with the known inhibitory effect of lead on a heme biosynthesis catalyst, aminolevulinate dehydratase (ALAD)^101^, and (lead induced) impairment of heme synthesis as a suggested potential trigger for multiple sclerosis^102^. Otherwise, we observed little evidence for causal roles of trace elements in the remaining selected health outcomes. True direct protective effects of non-essential or toxic trace elements seem implausible, although their effect could potentially be indirect, for example if the toxic trace elements influence essential trace elements in the body. Based on the PheWAS results and the known correlations and common biological pathways between several trace elements, the estimates were particularly vulnerable to misspecification of the primary phenotype and/or horizontal pleiotropy. Further, blood concentrations might not indicate other tissue or organ specific trace element concentrations. MR analyses can also be influenced by selection bias and competing risks, which might also explain apparent protective effects of toxic trace elements on late-life health outcomes. Few SNPs per instrument provided limited opportunities for reliable sensitivity analyses. Some estimates were precise around the null, but in general, the confidence intervals were wide and compatible with both moderate protection and harmful effects and were therefore uninformative. The low precision could be due to low case sample sizes in the outcome data and/or genetic instruments that had few SNPs with small to moderate effects on the exposures, potentially introducing weak instrument bias^103^. This was quantified by the low variance explained and low F-statistics of some of the instruments. These findings were based on the analyses of variation in trace elements concentrations in a general population, and we were therefore unable to estimate any effects of extreme trace element concentrations. The current results should be interpreted with caution, and future well-powered multivariable MR analyses might be helpful to discriminate between the effects of different trace elements. Further, as we only analyzed genetic associations with trace element concentrations measured in adults, it was not possible to evaluate any effects of trace elements specific to growth and development stages in newborns to adolescents^104^. Low sample sizes and lack of cohorts with both trace elements and health outcomes available also prevented us from exploring non-linear associations with MR.

This study has several limitations. Neither the HUNT nor the MoBa populations have generally been exposed to high levels of toxic trace elements, which makes these excellent populations for detecting genetic factors, but on the other hand many trace elements had a high proportion of measurements below the detection limit, thereby lowering the effective sample size. Further, trace elements are distributed differently in different organs, tissues, and body fluids^105–107^, and whole blood is therefore not the preferred tissue for monitoring the status of all trace elements. Some trace elements could also be influenced by evaporation, contamination from syringes or leakage of trace elements from glass vials. Differences between studies and populations also created known and potential limitations. Without access to individual level data in PIVUS, we were unable to harmonize all covariates. For example, the PIVUS association model included triglycerides and cholesterol as covariates, while the models used in HUNT and MoBa did not. Although HUNT, PIVUS and MoBa are all Scandinavian cohorts, the populations from which they have been sampled have clear differences, particularly in terms of birth year, age, and sex, and although the directions of effect were consistent across all three studies, the differences observed in effect size across these studies for a few SNPs could potentially be related to population-specific exposure patterns or factors related to pregnancy (MoBa). For example, fish intake might influence Swedes more than Norwegians, because there are indications that some toxic trace elements have accumulated more in fish caught in the shallow Bothnian Bay than along the deep Norwegian coastline ^108^. Further, pregnant women might change their diet and different generations could have been exposed to different levels of environmental pollution throughout their lives. The main limitation was that most of the trace elements were only measured in HUNT, and both low sample sizes and the lack of replication cohorts were general limitations. Further, trace element concentrations are associated with many different factors, and while we have controlled for some of these, there might still be unmeasured confounding by population stratification related to factors we have not controlled for, especially for rare variants. A particular limitation was the lack of detailed data on dietary habits. A particular limitation was the lack of detailed data on dietary habits. The heritability and genetic correlation estimates found using LD Score regression were limited by the potentially low polygenicity of the trace elements and genomic control (GC) correction of summary statistics, both which may contribute to heritability being underestimated. Additionally, using summary statistics from mixed models may contribute to overestimation. Finally, small sample sizes (and for some trace elements many measurements below the detection limit) resulted in highly imprecise estimates. The imprecision was especially large for the genetic correlations, which were further limited by heritability z-scores that were all below 4^109^.

In summary, we have identified novel genetic loci and replicated previously indicated loci for essential and non-essential trace elements. These highlighted interesting genes that may help establish biological pathways and mechanisms for uptake, distribution, metabolism, and excretion of trace elements in humans. MR analyses provided several indications of weak to moderate associations between trace elements and health outcomes, the most precise being a weak harmful effect of genetically determined circulating zinc on prostate cancer. However, generally imprecise MR estimates gave no convincing evidence for causal roles of genetically determined levels of other trace elements on health outcomes. Studies of populations with higher trace element exposure burdens or with larger samples are needed to investigate moderate and weak effects.

## Materials and methods

### Cohort Descriptions

#### HUNT

The HUNT Study^48, 49^ is a longitudinal population-based health study conducted in the county of Trøndelag, Norway. Data and samples have been collected through four cross-sectional surveys: HUNT1 (1984-86), HUNT2 (1995-97), HUNT3 (2006-08) and HUNT4 (2017-2019). Approximately 123 000 individuals (aged ≥ 20 years) have participated in one or more HUNT surveys. Approximately 88 000 individuals have been genotyped using one of three Illumina HumanCoreExome arrays: 12 v.1.0, 12 v.1.1 and 24 with custom content (UM HUNT Biobank v1.0). Sample and variant quality control (QC) was performed using standard practices and has been described elsewhere^110^. All variants were imputed from the HRC v1.1 reference panel^111^ merged with 2201 sequenced samples from HUNT, using Minimac3^112^.

Trace elements have been measured in whole blood samples collected in HUNT2 and HUNT3 using high-resolution inductively coupled plasma mass spectrometry (HR-ICP-MS) (Thermo Finnigan Element 2, Thermo Finnigan, Bremen, Germany) at three laboratories as part of previous studies. Here, we combined measurements of nine trace elements (As, Cd, Co, Cu, Pb, Mn, Hg, Se, Zn) in 930 samples (HUNT2) that have been analyzed by the National Institute of Occupational Health in Norway (STAMI), 53 trace elements (Al, Sb, As, Ba, Be, B, Br, Cd, Ca, Ce, Cs, Cl, Cr, Co, Cu, Ga, Ge, Au, Ho, In, Fe, La, Pb, Li, Mg, Mn, Hg, Mo, Nd, Ni, Nb, Pa, P, Pt, Pr, Re, Rh, Rb, Sm, Se, Si, Sr, S, Ta, Tb, Tl, Sn, W, U, V, Y, Zn, Zr) that have been measured in 757 samples (HUNT3) by one laboratory at NTNU (hereafter named NTNU1), and 30 trace elements (As, Be, Bi, B, Br, Cd, Ca, Cs, Cr, Cu, Ga, Au, In, Ir, Fe, Pb, Mg, Mn, Hg, Mo, Ni, Rb, Se, Ag, Sr, Tl, Th, Sn, W, Zn) that have been measured in 1539 samples (HUNT3) by a second laboratory at NTNU (hereafter named NTNU2). For 23 trace elements in HUNT3, some samples (maximum 30) had missing information on lab assignment, and we assigned these to NTNU2. There were no individuals with multiple measurements. Trace element measurements were returned to the HUNT Databank after sample and trace element QC at the respective laboratories. The samples analyzed by the STAMI laboratory were collected as part of a sub-study of iron status in women (selection criteria: female, age between 20 and 55 years old, non-pregnant, not blood donor in the past 2 years), the samples analyzed by HUNT1 were collected as part of a neuroimaging sub-study (selection criteria: age between 50 and 65, participation in previous HUNT surveys, exclusion criteria: MRI contraindications (pacemaker of the heart, clipped cerebral aneurysm, cochlear implants, severe claustrophobia, or body weight above 150 kg)), and 267 of the 1539 samples analyzed by NTNU2 were selected among type 2 diabetes cases as part of a diabetes related sub-study. Samples from the NTNU1 laboratory that were either below the detection limit or more than 10 times the median value had been removed and were not included in the current analysis. The specific reason for removal of these measurements was not available, therefore we excluded these measurements for our analysis. The proportion of unavailable samples per trace element from NTNU1 ranged from 0 to 69% (average 17%, median 2%). For samples from the NTNU2 laboratory, we replaced measurements below the detection limit with randomly generated numbers between 0 and the element-specific detection limit, because the true measurements were unavailable. The proportion of measurements below the detection limit from NTNU2 ranged from 0% to 97% (average 11%, median 0%). Details of sample collection, storage of samples, sample processing, quality control and ICP-MS analyses have been reported in detail elsewhere^8, 50, 51^.

#### MoBa

The Norwegian Mother, Father and Child Cohort Study (MoBa) is a population-based pregnancy cohort study conducted by the Norwegian Institute of Public Health. Participants were recruited from all over Norway from 1999-2008. The women consented to participation in 41% of the pregnancies. The cohort includes approximately 114 500 children, 95 200 mothers and 75 200 fathers^54^. The current study is based on version 12 of the quality-assured data files released for research in January 2019. The establishment of MoBa and initial data collection was based on a license from the Norwegian Data Protection Agency and approval from The Regional Committees for Medical and Health Research Ethics.

In short, pregnant women were recruited in their first trimester and invited to fill in three questionnaires during pregnancy, and to donate blood and urine samples at the time of ultrasound screening around gestational weeks 17–19 (mean 18.5). Blood samples were obtained from both parents during pregnancy and from mothers and children (umbilical cord) at birth. Follow-up is conducted through questionnaires and linkage to national health registries^54^. A total of 98 000 MoBa participants have been genotyped using one of three arrays: Illumina HumanCoreExome, Illumina Global Screening Array, or Illumina OmniExpress. In the current study, we used genetic data collected as part of the Better Health by Harvesting Biobanks (HARVEST) study released by MoBa Genetics v.1.0 (https://github.com/folkehelseinstituttet/mobagen/wiki/MoBaGenetics1.0). The sample and variant quality control in MoBa has been described elsewhere^113^. All variants were imputed from the HRC reference panel v1.1 at the Sanger Imputation Service.

The Norwegian Environmental Biobank is a sub-study within MoBa established with the aim of biomonitoring nutrients and environmental contaminants in MoBa participants. The study included 2999 pregnant women with available genetic data who had donated blod and urine samples and had responded to questionnaires 1-6 in MoBa^7, 53^. A total of 11 trace elements (As, Cd, Co, Cu, Mn, Mo, Pb, Se, Tl, Zn, Hg), were measured in whole blood donated by the women in gestational week 18 at the department of laboratory medicine at Lund University (Sweden). All trace elements except Hg were analyzed using an ICP-MS (iCAP Q, Thermo Fisher Scientific, Bremen, Germany). Total Hg was determined in acid-digested samples by cold vapor atomic fluorescence spectrophotometry (Sandborgh-Englund 1998). During the analysis campaign, the laboratory participated in the German External Quality Assessment Scheme (G-EQAS), with good agreement between obtained element concentrations in quality control samples used and expected values. Details of sample collection, storage of samples, sample processing and ICP-MS analyses have been reported in detail elsewhere^7^.

#### Association analyses

We performed genome-wide association analyses of 59 trace elements measured in up to 2819 individuals in HUNT and 11 trace elements measured in 2812 individuals in MoBa. For 48 trace elements in HUNT (Al, Sb, As, Ba, Be, Bi, B, Br, Cd, Ca, Ce, Cs, Cl, Co, Cu, Ga, Ge, Au, Ho, In, Ir, Fe, La, Pb, Li, Mg, Mn, Hg, Mo, Nd, Nb, Pa, P, Rh, Rb, Sm, Se, Ag, Sr, S, Ta, Tb, Tl, Sn, W, U, Y, Zn) and 9 trace elements in MoBa (Cd, Co, Cu, Hg, Mn, Mo, Pb, Se, Tl, Zn), we used a linear mixed model regression under an additive genetic model for each variant as implemented in BOLT-LMM v.2.3.4^114^, thereby controlling for relatedness between study participants. We performed association analyses of 9 trace elements in HUNT (Cr, Ni, Pt, Pr, Re, Si, Th, V, Zr) and 2 trace elements in MoBa (As, Mo) in unrelated individuals using PLINK 2.0^115^, because BOLT-LMM was unable to estimate the trace element heritability. In total, human GWASs had not been previously published for 39 trace elements (Sb, Ba, Be, B, Br, Bi, Ce, Cs, Ga, Ge, Au, Ho, In, Ir, La, Li, Nd, Nb, Pd, Pt, Pr, Re, Rh, Rb, Sm, Si, Ag, Sr, S, Ta, Tb, Tl, Th, Sn, W, U, V, Y, Zr). Distributions, sample size and proportion of measurements below the detection limit per study are given for each trace element in Supplementary Table 1.

Prior to analysis, we applied rank-based inverse normal transformation of the trace element concentrations after adjusting for age and sex (HUNT) using linear regression. Age, and the first ten genetic principal components (PCs) of ancestry were included as covariates in all the analyses. Sex, genotyping batch, geographical region (coast, town/fjord, or inland/mountain) and analysis laboratory (NTNU1, NTNU2, STAMI) were included as additional covariates in HUNT where appropriate. We performed genomic control correction of all GWAS results with an inflation factor λ > 1 (calculated from variants with MAF ≥ 0.01).

Variants with a minor allele count < 10 or an imputation R^2^ < 0.3 were excluded from the analyses. After visual inspection of the Manhattan and quantile-quantile plots, we excluded the full set of results for Bi and Th due to excessive inflation of the p-values (Supplementary Figures 42-45).

We used METAL^116^ to perform fixed-effect inverse variance weighted GWA meta-analysis of 14 trace elements (Al, As, Cd, Cr, Co, Cu, Pb, Mn, Hg, Mo, Se, Tl, Zn) analyzed in at least two of three studies: the HUNT, MoBa and/or publicly available summary statistics from the PIVUS study^15^, where 11 trace elements (Al, Cd, Co, Cu, Cr, Hg, Mn, Mo, Ni, Pb, Zn) have been measured in 949 seniors from Uppsala, Sweden, and rank inverse normalized trace element concentrations had been tested for association using a linear regression under an additive model, adjusting for triglycerides, cholesterol, gender and two principal components of ancestry. Age was not included as covariate because the PIVUS participants were of the same age.

#### Definition of associated variants, associated loci, and locus novelty

We considered genetic variants with p-values < 5×10^−8^ to be statistically significant at a genome-wide significance level and defined these as associated with the given trace element. Genetic loci were defined as 500 kilobasepairs to each side of genome-wide significant variants in the same region. A locus was classified as novel for a given trace element if it had not been reported for the respective trace element before. Previously published loci were identified with a literature search and a look-up in the GWAS catalog^117^. Index SNPs were identified as the genetic variant with the lowest p-value in each locus. We used PLINK v.1.9^115^ to identify variants in high LD (correlation r^2^>0.8) with the index variants, based on a reference panel of 5000 unrelated individuals in HUNT. The index variants and variants in high LD with the index variants were annotated using ANNOVAR^118^.

#### Sensitivity analyses

For trace elements where tobacco smoke (Cd, As, Pb, Cu)^119^, wine (Pb)^120^ or fish (As, Se)^121^ is a major source of human intake, or where alcohol is thought to regulate uptake or metabolism (Fe, Mn, Zn)^122–124^, we repeated the association analyses of the associated loci in HUNT, including smoking status (self-reported, never versus ever smokers (including ex-smokers, occasionally and daily smokers)), frequency of fat fish intake (self-reported) and units of alcohol per week (self-reported) as covariates, respectively (Supplementary Table 4). Further, since some of the analyzed samples were selected from type 2 diabetes cases, and type 2 diabetes is associated with hypomagnesemia^89^, we repeated the association analyses of magnesium including diabetes status (excluding type 1 diabetes) as covariate. Diabetes cases were defined as either any non-type 1 diabetes (self-reported) and/or fasting serum glucose ≥ 7.0 mmol/liter and/or serum glucose ≥ 11.0 mmol/liter 2 hours after first having fasted overnight and then consumed 75 grams of glucose dissolved in ∼3 dl water.

#### Heritability estimation and genetic and phenotypic correlation between trace elements

We used LD Score regression^109, 125^ to estimate the narrow-sense SNP heritability (V_g_/V_p_ ± 1SE, where V_g_ is the variance explained by the SNPs and V_p_ is the total phenotypic variance) of 11 trace elements from the GWA meta-analysis summary level SNP results, using LD scores estimated from individuals of European ancestry in the HUNT population. The analysis was performed in trace elements with a meta-analysis sample size > 5000, as recommended for the LD Score regression software. Each set of summary statistics were restricted to well imputed SNPs in HapMap3. Further, we used LD Score regression to estimate the genetic correlation between all pairs of 10 trace elements with an estimated SNP heritability > 0 and a sample size > 5000 in the GWA meta-analysis. The cadmium-mercury and molybdenum-cobalt correlations were excluded because the genetic estimates were out of bounds (higher than 1.0 or lower than −1.0). Additionally, we estimated the phenotypic correlation between the same pairs of trace elements in HUNT using Spearman rank correlation. Prior to the phenotypic correlations, the trace elements were corrected for median concentration per lab and log2 transformed.

#### Phenome-wide association tests (PheWAS)

We tested for associations of 34 common and low-frequency (MAF>0.5%) trace element index variants (21 from meta-analysis and 13 from GWAS in HUNT) with 1326 phecodes, 167 continuous traits and 30 biomarkers in participants of white British ancestry in the UK Biobank, using publicly available summary statistics (https://pan.ukbb.broadinstitute.org/). Four variants (rs146233512 [gold], rs763064690 [indium], rs78394934 [iron], rs927502065 [tungsten]) were excluded because they were not tested in the UK Biobank. To correct for the total number of tests (n=51782), we used a Bonferroni corrected p-value significance threshold of 9.7×10^−7^.

#### Mendelian randomization (MR) of individual trace elements on selected health related outcomes

To explore potential causal associations of trace elements on selected outcomes, we used two-sample MR: We applied the inverse-variance weighted (IVW) method for trace elements with multiple index SNPs, and the Wald ratio method for trace elements with only one index SNP, as implemented in the TwoSampleMR^126^ and MRInstruments^127^ packages in R v3.6.3 and R v4.0.5. The exposures were selected based on robust genetic associations with trace elements in the current study, i.e. meta-analyzed index variants and the common index variant in the locus strongly associated with Sr in HUNT. The outcomes were selected based the availability of instrument summary statistics for a priori outcomes of interest highlighted in previous literature: Alzheimer’s disease (Mn, Pb, As, Cd, Cu, Se and Zn)^20, 21, 25–28, 128^, Parkinson’s disease (Pb, As, Cd, Cu, Mn, Se and Zn)^20, 29–31^, multiple sclerosis (Zn, Mn, Se, Pb, As and Cd)^22–24^, autoimmune thyroid disease (Se)^33^, hypothyroidism (Se)^33^, osteoporosis (Cu)^44, 46^, bone mineral density (Cu, Cd, Mn, Zn and Sr)^45–47^, bone fracture (Sr)^47^, rheumatoid arthritis (Cd and Zn)^34, 35^, type 2 diabetes (Cu, Mn, Se and Zn)^38–40^, colorectal cancer (Zn and Se)^42, 43^ and prostate cancer (Se, Zn)^41, 43^. We used SNP-exposure associations from the GWA meta-analysis (Cu, Zn, Mn, Se, Pb, As, Cd) or GWAS in HUNT (Sr) of each trace element (Supplementary Table 2), and SNP-outcome associations were collected from independent genome-wide summary level data^129–139^ (Supplementary Table 13). For the SNP associations with multiple sclerosis, we calculated the beta coefficient as the natural logarithm of the OR, and the standard error as 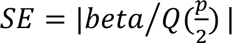, where | | denotes absolute values, p denotes the p-value and *Q* denotes the inverse standard normal distribution. Outcomes obtained from https://pan.ukbb.broadinstitute.org (Pan-UKB team, 2020) were defined as: hypothyroidism (phecode: 244), and osteoporosis (phecode: 743.1). We estimated the variance explained and F-statistic for the instruments from a linear regression of the trace element concentrations versus the sum of estimated allele counts (dosages) for the trace element increasing alleles in HUNT (Supplementary Table 13). Except for manganese, the trace element instruments consisted of fewer than 4 SNPs, which provided limited opportunities for reliable sensitivity analyses.

#### Ethics

The current study has been approved by the Norwegian Data Protection Authority and approved by the Regional Ethics Committee for Medical and Health Research Ethics in Northern Norway (REK Reference Number: 51604). All MoBa and HUNT participants have signed informed written consent. The MoBa cohort is currently regulated by the Norwegian Health Registry Act.

## Data Availability

Summary level data supporting the findings are available in the Supplementary materials and upon request. The consent given by the participants in HUNT and MoBa does not open for storage of individual level data in repositories or journals. Researchers associated with Norwegian research institutes can apply for the use of HUNT data. Researchers from other countries may apply if collaborating with a Norwegian Principal Investigator. Information for data access can be found at https://www.ntnu.edu/hunt/data. Researchers who want access to MoBa data sets for replication should apply to helsedata.no. Access to data from either HUNT or MoBa requires approval from The Regional Committees for Medical and Health Research Ethics in Norway.

## Acknowledgments

The Trøndelag Health Study (The HUNT Study) is a collaboration between HUNT Research Center (Faculty of Medicine and Health Sciences, NTNU, Norwegian University of Science and Technology), Trøndelag County Council, Central Norway Regional Health Authority, and the Norwegian Institute of Public Health. The genotyping in HUNT was financed by the National Institutes of Health; University of Michigan; the Research Council of Norway; the Liaison Committee for Education, Research and Innovation in Central Norway; and the Joint Research Committee between St Olav’s hospital and the Faculty of Medicine and Health Sciences, NTNU. The genetic investigations of the HUNT Study are a collaboration between researchers from the K.G. Jebsen Center for Genetic Epidemiology, NTNU, and the University of Michigan Medical School and the University of Michigan School of Public Health. The K.G. Jebsen Center for Genetic Epidemiology is financed by Stiftelsen Kristian Gerhard Jebsen; Faculty of Medicine and Health Sciences, NTNU, Norway. We thank HUNT participants for donating their time, samples, and information to help others. The Norwegian Mother, Father and Child Cohort Study (MoBa) is supported by the Norwegian Ministry of Health and Care Services and the Ministry of Education and Research. We are grateful to all the participating families in Norway who take part in this on-going cohort study. We thank the Norwegian Institute of Public Health (NIPH) for generating high-quality genomic data. This research is part of the HARVEST collaboration, supported by the Research Council of Norway (#229624). We also thank the NORMENT Centre for providing genotype data, funded by the Research Council of Norway (#223273), South East Norway Health Authorities and Stiftelsen Kristian Gerhard Jebsen. We further thank the Center for Diabetes Research, the University of Bergen for providing genotype data and performing quality control and imputation of the data funded by the ERC AdG project SELECTionPREDISPOSED, Stiftelsen Kristian Gerhard Jebsen, Trond Mohn Foundation, the Research Council of Norway, the Novo Nordisk Foundation, the University of Bergen, and the Western Norway Health Authorities. The NIPH has contributed to funding of the Norwegian Environmental Biobank (NEB). The laboratory measurements have partly been funded by the Research Council of Norway through research projects (275903 and 268465), and the human biomonitoring project HBM4EU, funded by the European Union’s Horizon 2020 research and innovation programme under grant agreement No 733032.

## Author contributions

M.R.M. analyzed the data and wrote the first draft of the manuscript. B.M.B., K.H. and C.J.W. conceived and designed the study. A.F.H., B.N.W., L.F.T, H.R. and L.B. contributed to analyses. A.S., T.S., T.H.N. and T.P.F gave advice on phenotypes and phenotype definitions. K.H., C.W., P.M., P.R.N., O.A.A, A.L.B., T.S., A.S. and T.P.F. contributed to data acquisition. M.R.M., A.F.H., I.S., W.Z., J.Z, L.F., D.M.E., N.M.W., T.H.N., B.O.Å., T.P.F., C.J.W., K.H. and B.M.B. interpreted the results. All authors revised the final version of the paper.

