## Supplementary Figures for "New insights into the genetic etiology of 57 essential and non-essential trace elements in humans"

**Supplementary Figures 1-14:** Manhattan and quantile-quantile (QQ) plots for trace elements with significant findings in GWA meta-analysis

**Supplementary Figures 15-38:** Manhattan and QQ plots for trace elements with significant findings in GWAS in HUNT

**Supplementary Figure 39:** Pairwise phenotypic and genetic correlations of meta-analyzed trace elements

**Supplementary Table 7:** Pairwise phenotypic and genetic correlations of meta-analysed trace elements

**Supplementary Figure 40**: Phenome-wide associations with GWA meta-analysis index variants

**Supplementary Figure 41**: Phenome-wide associations with GWAS index variants in HUNT

**Supplementary Figure 42-45:** Manhattan and QQ plots for trace elements that were excluded due to inflated test statistics

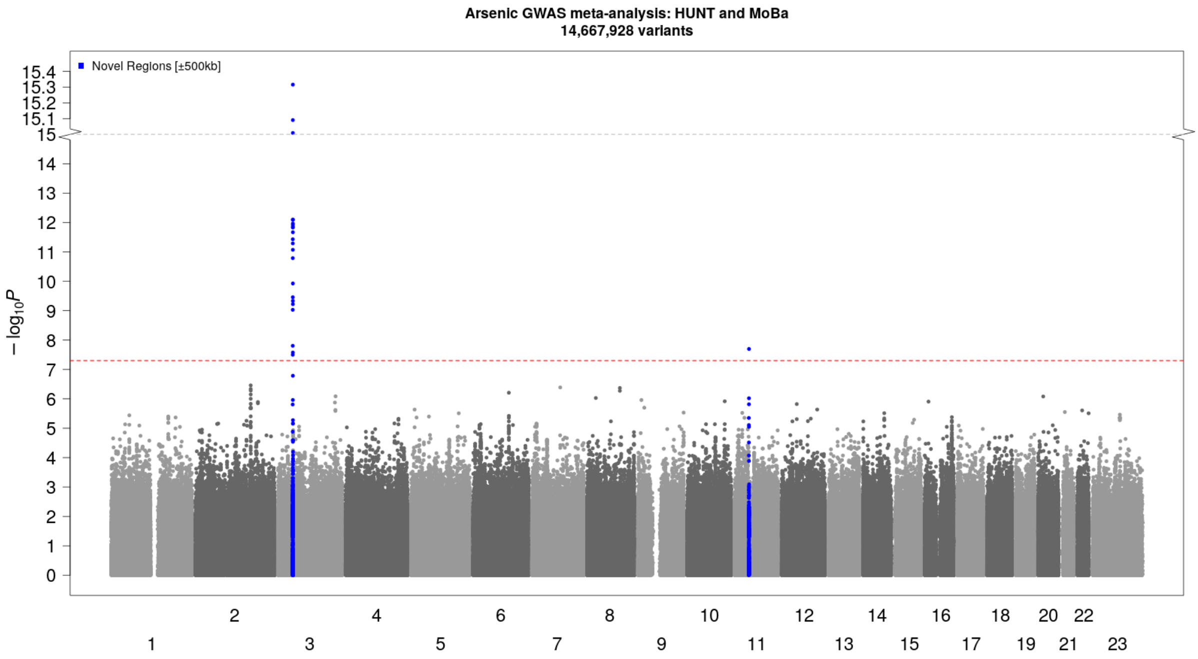


***Supplementary Figure 1: Manhattan plot [arsenic]:*** *Novel (blue) genetic loci (index variant ±500 kilobase pairs) associated with whole blood arsenic concentrations at genome-wide significance (p-value<5×10*$^{-8}$*, threshold indicated by red dotted line) in HUNT and MoBa. Genetic variants plotted according to chromosome and position (x-axis) and the -log10(p-value) for the variant-arsenic level association (y-axis). Sample size N=5591. HUNT=The Trøndelag Health Study. MoBa=The Norwegian Mother, Father, and Child Cohort Study.*

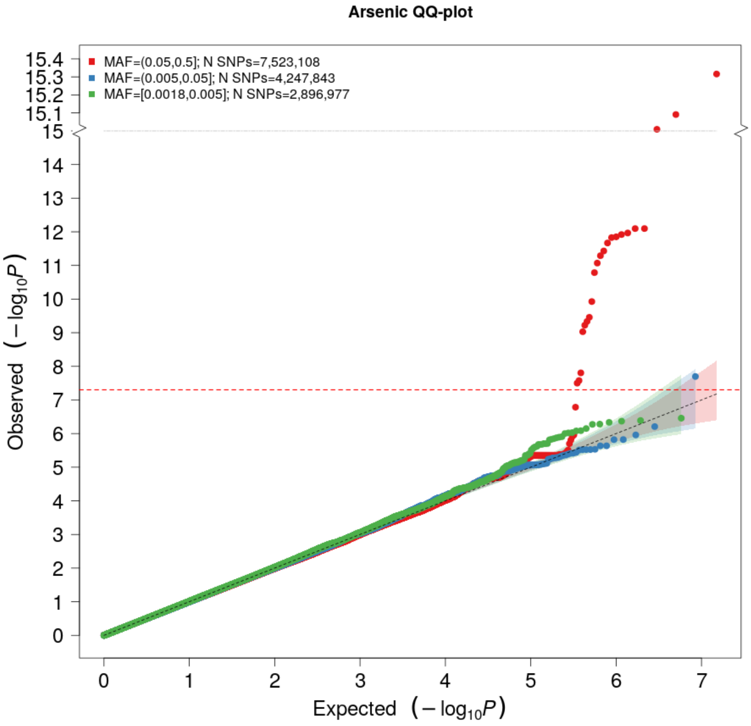


***Supplementary Figure 2: QQ plot [Arsenic]:*** *The expected -log10(p-value) given no true associations are given on the x-axis, and the observed -log10(p-value) from the GWA meta-analysis of arsenic concentrations in HUNT and MoBa are given on the y-axis, stratified by minor allele frequency (MAF) categories: Common (MAF>0.05) variants (red), low-frequency (0.005<MAF<0.05) variants (blue) and rare (MAF<0.005) variants (green). HUNT=The Trøndelag Health Study. MoBa=The Norwegian Mother, Father, and Child Cohort Study.*

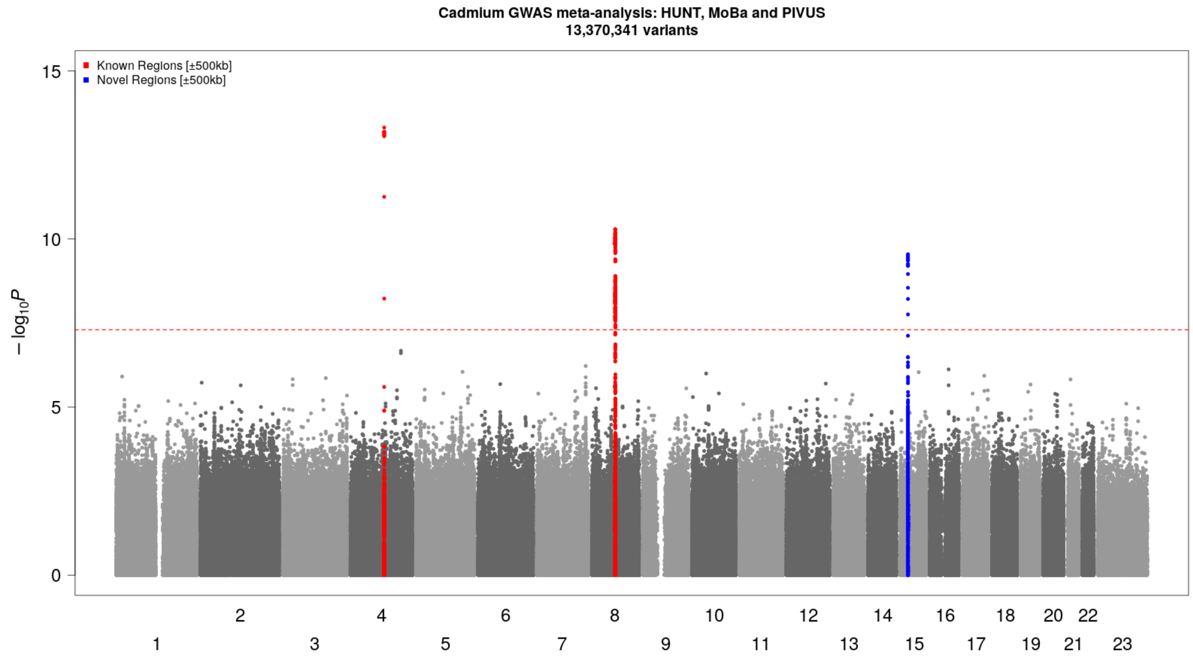


***Supplementary Figure 3: Manhattan plot [cadmium]:*** *Novel (blue) and known (red) genetic loci (index variant ±500 kilobase pairs) associated with whole blood cadmium concentrations at genome-wide significance (p-value<$5 \times 10^{-8}$, threshold indicated by red dotted line) in HUNT, MoBa and PIVUS. Genetic variants plotted according to chromosome and position (x-axis) and the -log10(p-value) for the variant-cadmium level association (y-axis). Sample size N=6542. HUNT=The Trøndelag Health Study. MoBa=The Norwegian Mother, Father, and Child Cohort Study. PIVUS=The Prospective Investigations of the Vasculature in Uppsala Seniors study.*

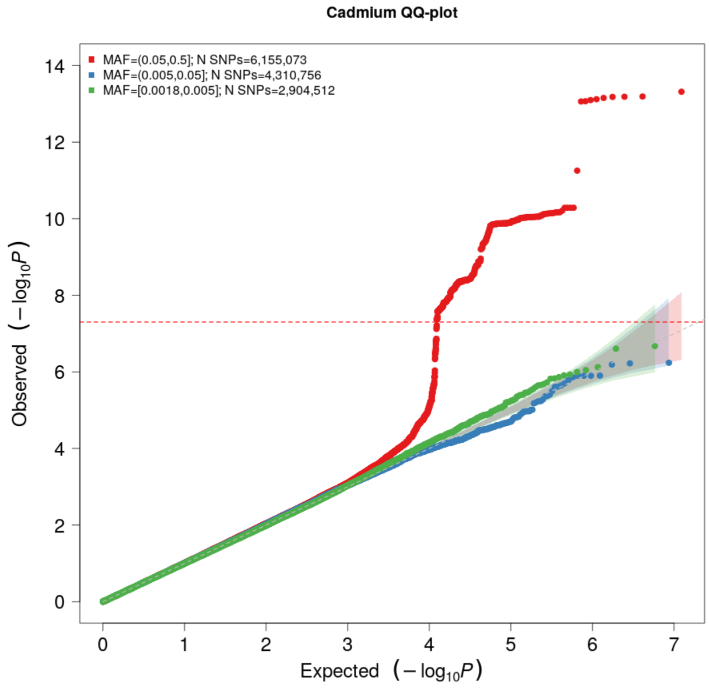


***Supplementary Figure 4: QQ plot [cadmium]:*** *The expected -log10(p-value) given no true associations are given on the x-axis, and the observed -log10(p-value) from the GWA meta-analysis of cadmium concentrations in HUNT, MoBa and PIVUS are given on the y-axis, stratified by minor allele frequency (MAF) categories: Common (MAF>0.05) variants (red), low-frequency (0.005<MAF<0.05) variants (blue) and rare (MAF<0.005) variants (green). HUNT=The Trøndelag Health Study. MoBa=The Norwegian Mother, Father, and Child Cohort Study. PIVUS=The Prospective Investigations of the Vasculature in Uppsala Seniors study.*

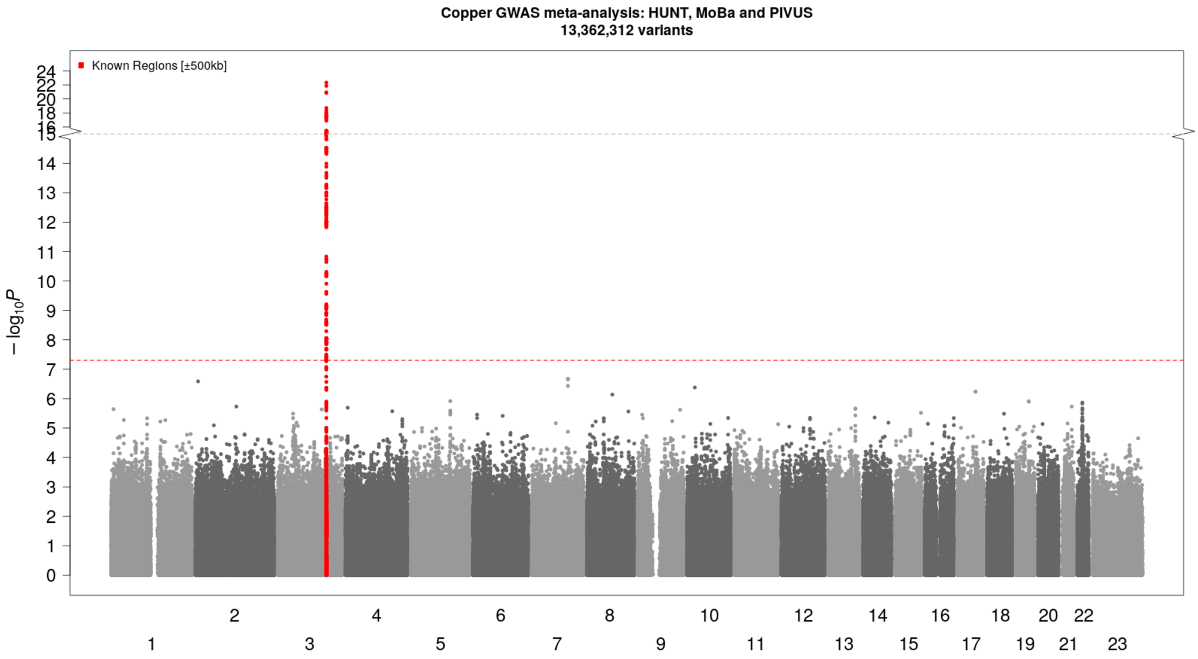


***Supplementary Figure 5: Manhattan plot [copper]:*** *Known (red) genetic locus (index variant ±500 kilobase pairs) associated with whole blood copper concentrations at genome-wide significance (p-value<5×10⁻⁸, threshold indicated by red dotted line) in HUNT, MoBa and PIVUS. Genetic variants plotted according to chromosome and position (x-axis) and the -log10(p-value) for the variant-copper level association (y-axis). The red dotted line indicates the genome-wide significance threshold. Sample size N=6564. HUNT=The Trøndelag Health Study. MoBa=The Norwegian Mother, Father, and Child Cohort Study. PIVUS=The Prospective Investigations of the Vasculature in Uppsala Seniors study.*

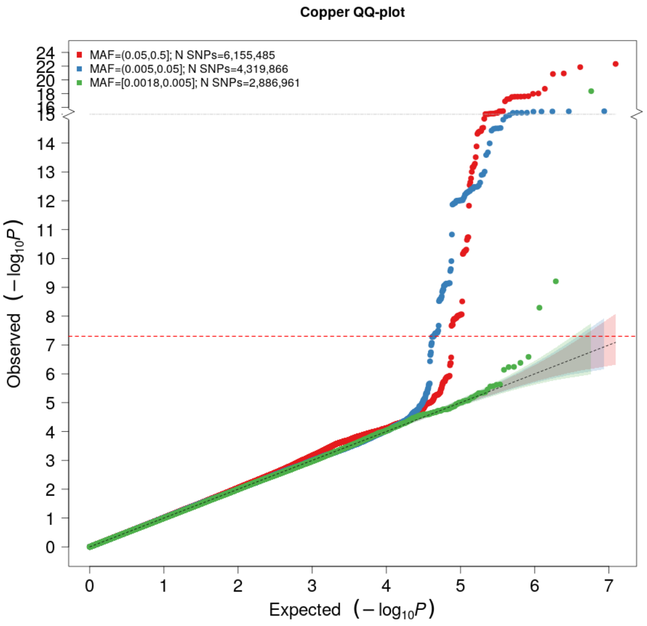


***Supplementary Figure 6: QQ plot [copper]:*** *The expected -log10(p-value) given no true associations are given on the x-axis, and the observed -log10(p-value) from the GWA meta-analysis of copper in HUNT, MoBa and PIVUS are given on the y-axis, stratified by minor allele frequency (MAF) categories: Common (MAF>0.05) variants (red), low-frequency (0.005<MAF<0.05) variants (blue) and rare (MAF<0.005) variants (green). HUNT=The Trøndelag Health Study. MoBa=The Norwegian Mother, Father, and Child Cohort Study. PIVUS=The Prospective Investigations of the Vasculature in Uppsala Seniors study.*

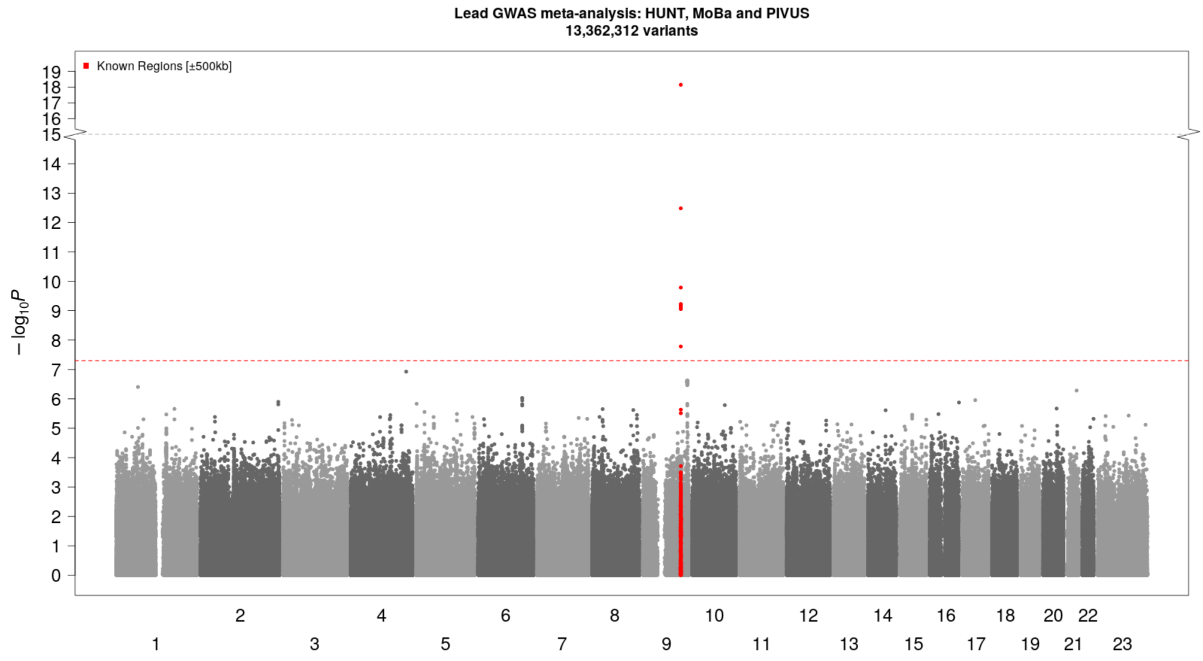


***Supplementary Figure 7: Manhattan plot [lead]:*** *Known (red) genetic locus (index variant ±500 kilobase pairs) associated with whole blood lead concentrations at genome wide significance (p-value<5×10⁻⁸, threshold indicated by red dotted line) in HUNT, MoBa and PIVUS. Genetic variants plotted according to chromosome and position (x-axis) and the -log10(p-value) for the variant-lead level association (y-axis). Sample size N=6564. HUNT=The Trøndelag Health Study. MoBa=The Norwegian Mother, Father, and Child Cohort Study. PIVUS=The Prospective Investigations of the Vasculature in Uppsala Seniors study.*

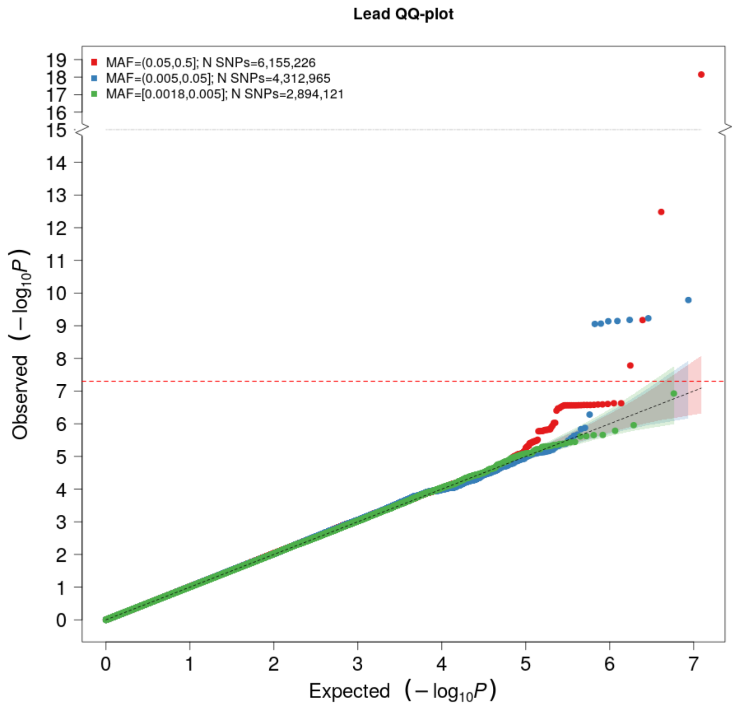


***Supplementary Figure 8: QQ plot [lead]:*** *The expected -log10(p-value) given no true associations are given on the x-axis, and the observed -log10(p-value) from the GWA meta-analysis of lead concentrations in HUNT, MoBa and PIVUS are given on the y-axis, stratified by minor allele frequency (MAF) categories: Common (MAF>0.05) variants (red), low-frequency (0.005<MAF<0.05) variants (blue) and rare (MAF<0.005) variants (green). HUNT=The Trøndelag Health Study. MoBa=The Norwegian Mother, Father, and Child Cohort Study. PIVUS=The Prospective Investigations of the Vasculature in Uppsala Seniors study.*

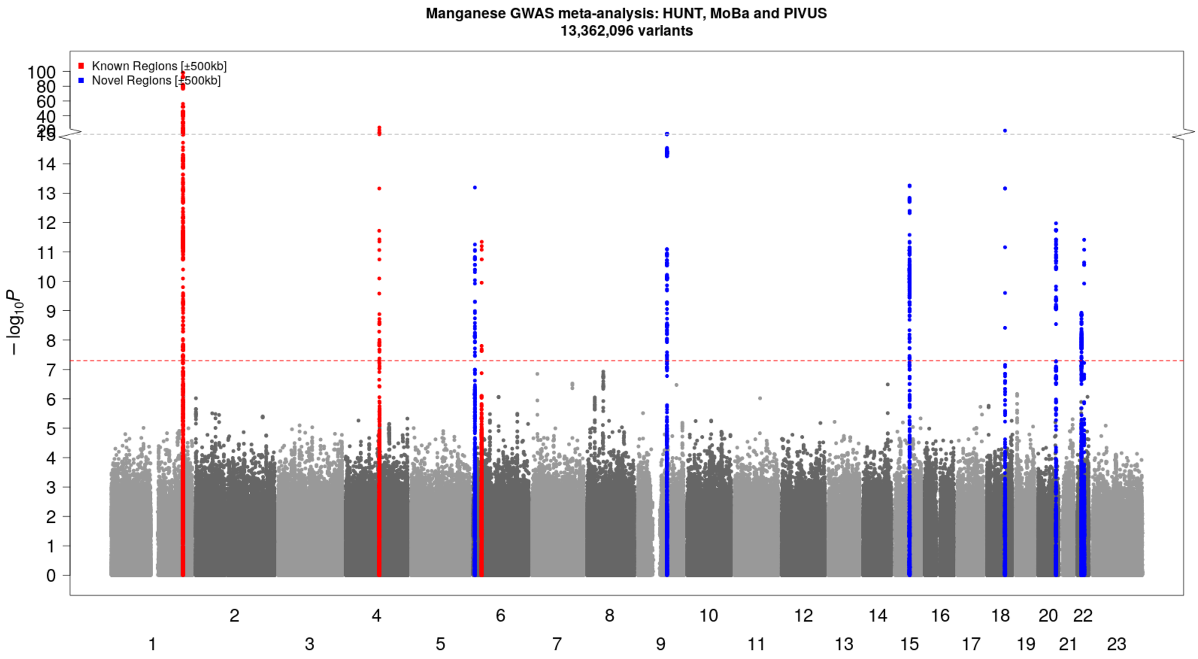


***Supplementary Figure 9: Manhattan plot [manganese]:*** *Novel (blue) and known (red) genetic loci (index variant ±500 kilobase pairs) associated with whole blood manganese concentrations at genome-wide significance (p-value<$5\times10^{-8}$, threshold indicated by red dotted line) in HUNT, MoBa and PIVUS. Genetic variants are plotted according to chromosome and position (x-axis) and the -log10(p-value) for the variant-manganese level association (y-axis). Sample size N=6564. HUNT=The Trøndelag Health Study. MoBa=The Norwegian Mother, Father, and Child Cohort Study. PIVUS=The Prospective Investigations of the Vasculature in Uppsala Seniors study.*

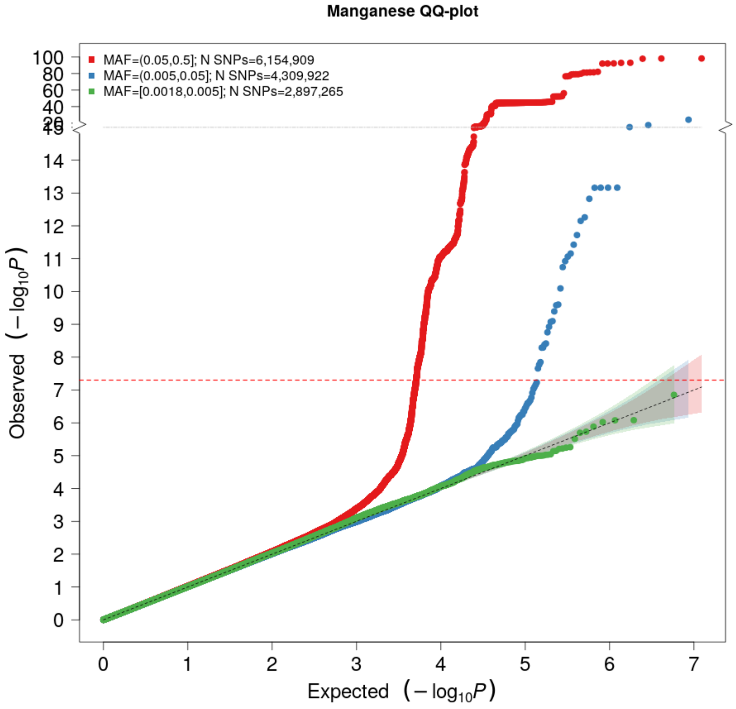


***Supplementary Figure 10: QQ plot [manganese]:*** *The expected -log10(p-value) given no true associations are given on the x-axis, and the observed -log10(p-value) from the GWA meta-analysis of manganese concentrations in HUNT, MoBa and PIVUS are given on the y-axis, stratified by minor allele frequency (MAF) categories: Common (MAF>0.05) variants (red), low-frequency (0.005<MAF<0.05) variants (blue) and rare (MAF<0.005) variants (green). HUNT=The Trøndelag Health Study. MoBa=The Norwegian Mother, Father, and Child Cohort Study. PIVUS=The Prospective Investigations of the Vasculature in Uppsala Seniors study.*

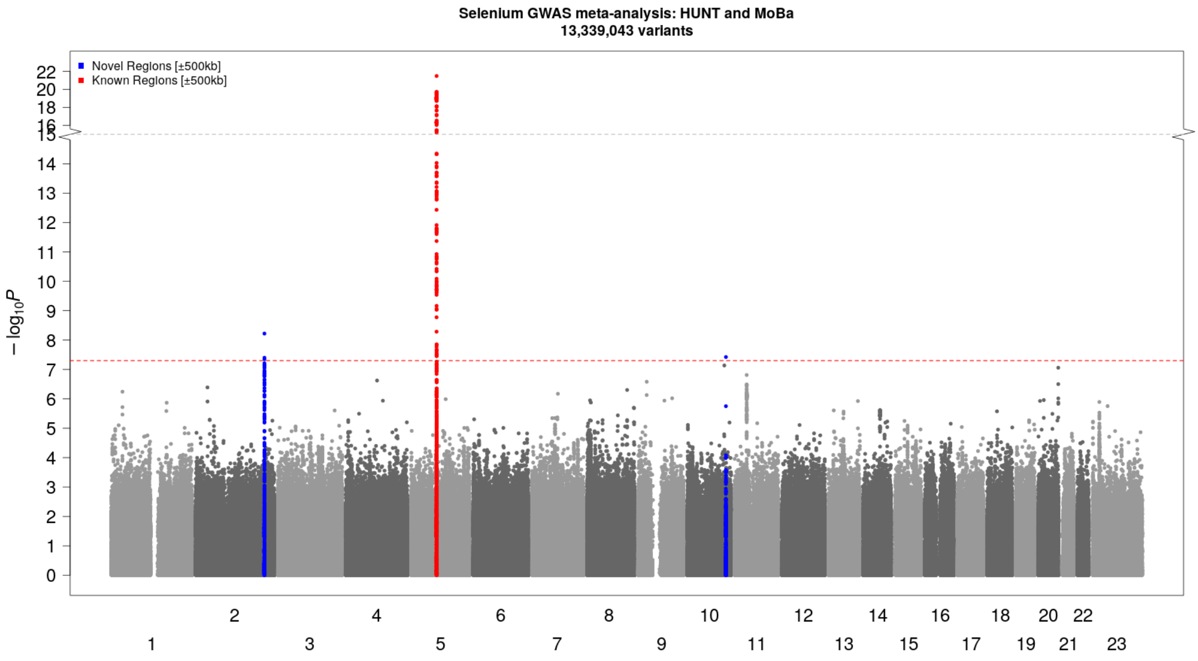


***Supplementary Figure 11: Manhattan plot [selenium].*** *Novel (blue) and known (red) genetic loci (index variant ±500 kilobase pairs) associated with whole blood selenium concentrations at genome-wide significance (p-value<5×10⁻⁸, threshold indicated by red dotted line) in HUNT and MoBa. Genetic variants plotted according to chromosome and position (x-axis) and the -log10(p-value) for the variant-selenium association (y-axis). Note: What appears to be a single SNP association on chromosome 10, is a genetic locus where other variants correlated ($R^2$>0.8) with the index variant in HUNT did not replicate in MoBa. However, the index variant was imputed only in HUNT, and is therefore visible in the figure. Sample size N=5615. HUNT=The Trøndelag Health Study. MoBa=The Norwegian Mother, Father, and Child Cohort Study.*

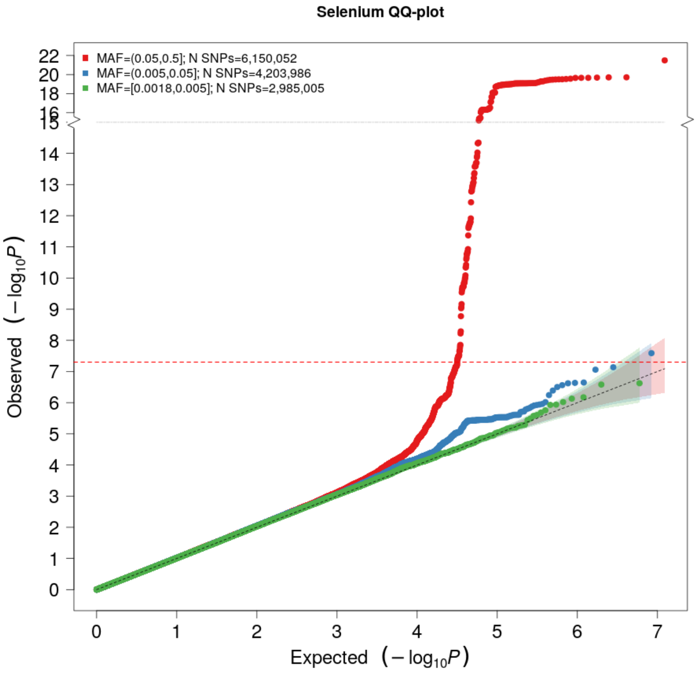


***Supplementary Figure 12: QQ plot [selenium]:*** *The expected -log10(p-value) given no true associations are given on the x-axis, and the observed -log10(p-value) from the GWA meta-analysis of selenium concentrations in HUNT and MoBa are given on the y-axis, stratified by minor allele frequency (MAF) categories: Common (MAF>0.05) variants (red), low-frequency (0.005<MAF<0.05) variants (blue) and rare (MAF<0.005) variants (green). HUNT=The Trøndelag Health Study. MoBa=The Norwegian Mother, Father, and Child Cohort Study.*

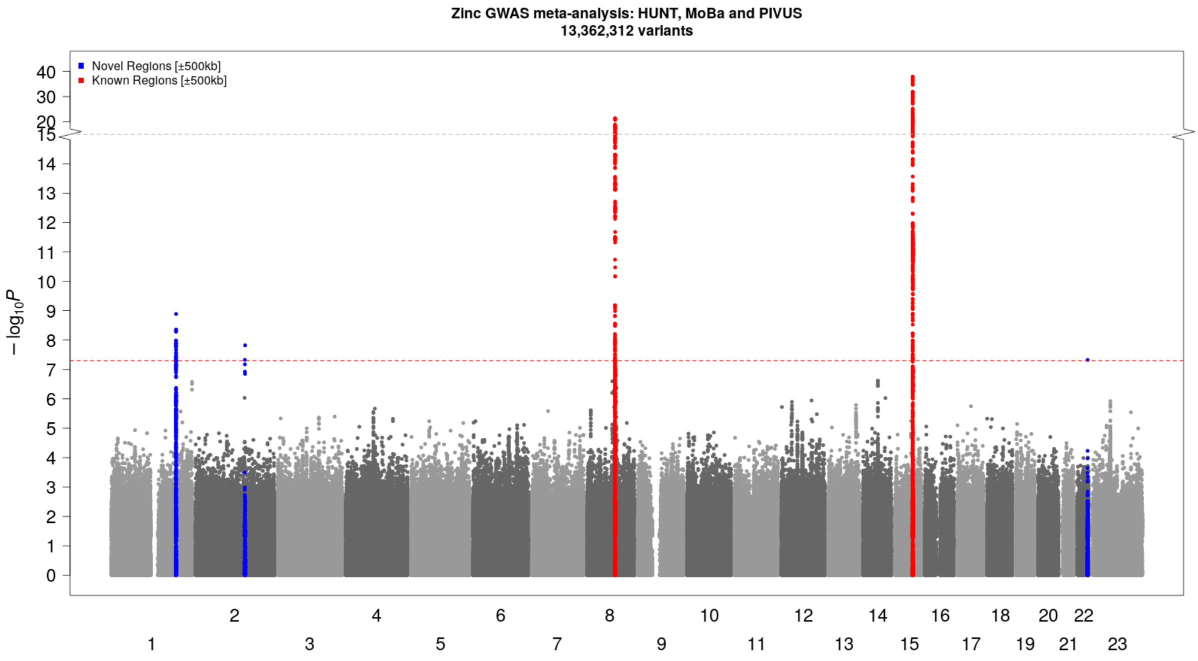


***Supplementary Figure 13: Manhattan plot [zinc]:*** *Novel (blue) and known (red) genetic loci (index variant ±500 kilobase pairs) associated with whole blood zinc concentrations at genome-wide significance (p-value<$5\times10^{-8}$, threshold indicated by red dotted line) in HUNT, MoBa and PIVUS. Genetic variants plotted according to chromosome and position (x-axis) and the -log10(p-value) for the variant-zinc level association (y-axis). Sample size N=6564. HUNT=The Trøndelag Health Study. MoBa=The Norwegian Mother, Father, and Child Cohort Study. PIVUS=The Prospective Investigations of the Vasculature in Uppsala Seniors study.*

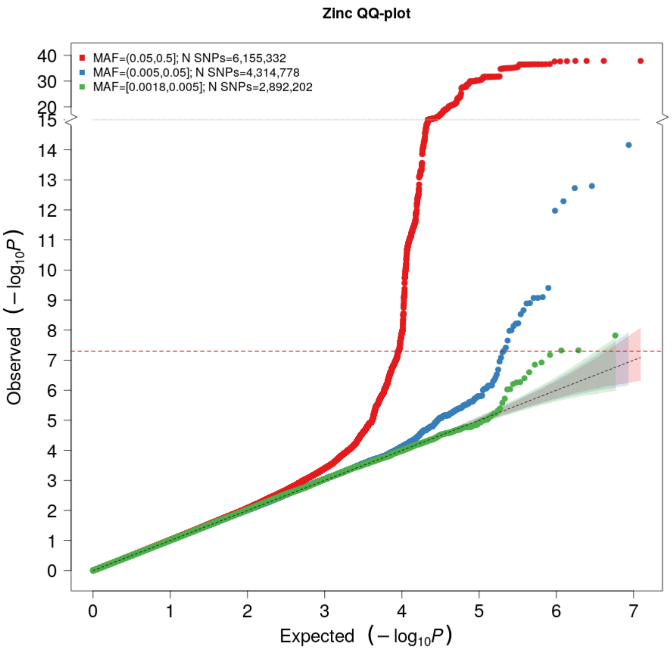


***Supplementary Figure 14: QQ plot [zinc].*** *The expected -log10(p-value) given no true associations are given on the x-axis, and the observed -log10(p-value) from the GWA meta-analysis of whole blood zinc concentrations in HUNT, MoBa and PIVUS are given on the y-axis, stratified by minor allele frequency (MAF) categories: Common (MAF>0.05) variants (red), low-frequency (0.005<MAF<0.05) variants (blue) and rare (MAF<0.005) variants (green). HUNT=The Trøndelag Health Study. MoBa=The Norwegian Mother, Father, and Child Cohort Study. PIVUS=The Prospective Investigations of the Vasculature in Uppsala Seniors study.*

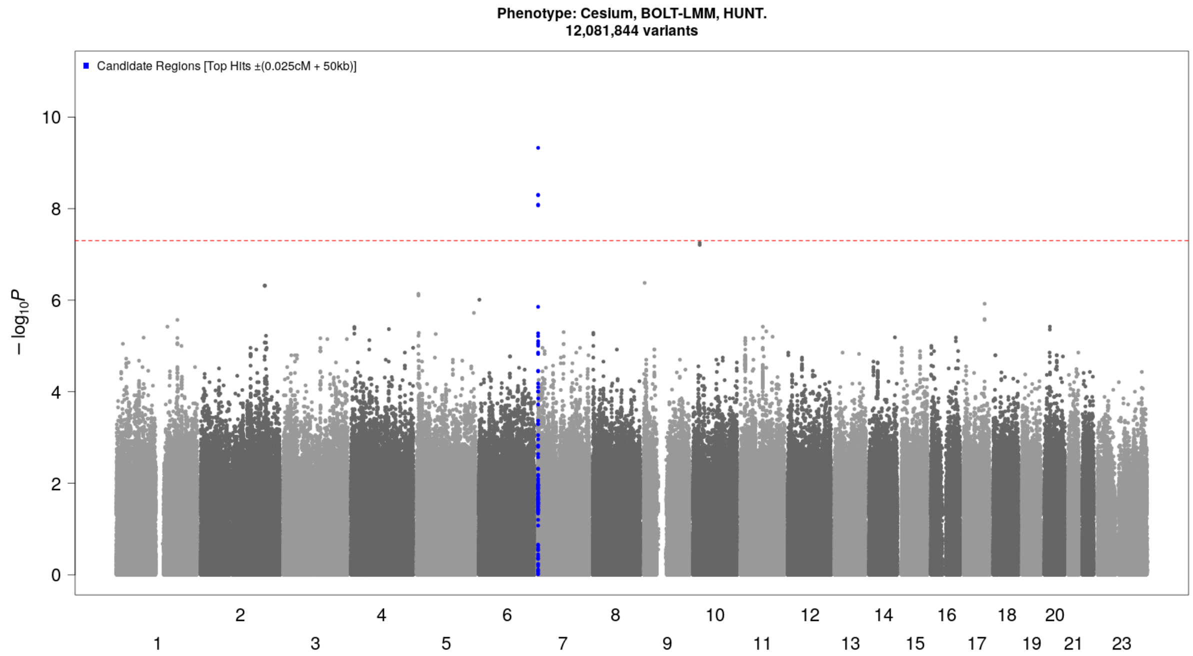


***Supplementary Figure 15: Manhattan plot [cesium]:*** *Novel (candidate) genetic loci (index variant ±500 kilobase pairs) associated with whole blood cesium concentrations at genome-wide significance (p-value<5×10^-8^, threshold indicated by red dotted line) in HUNT. Genetic variants plotted according to chromosome and position (x-axis) and the -log10(p-value) for the variant-cesium level association (y-axis). Sample size N=2197. HUNT=The Trøndelag Health Study.*

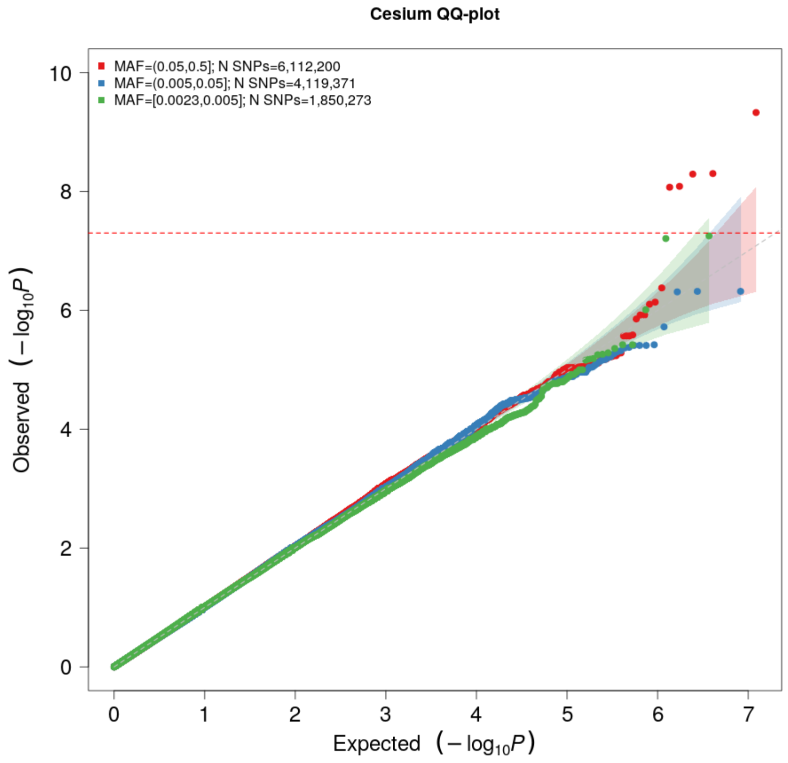


***Supplementary Figure 16: QQ plot [cesium].*** *The expected -log10(p-value) given no true associations are given on the x-axis, and the observed -log10(p-value) from the GWA meta-analysis of whole blood cesium concentrations in HUNT are given on the y-axis, stratified by minor allele frequency (MAF) categories: Common (MAF>0.05) variants (red), low-frequency (0.005<MAF<0.05) variants (blue) and rare (MAF<0.005) variants (green). HUNT=The Trøndelag Health Study.*

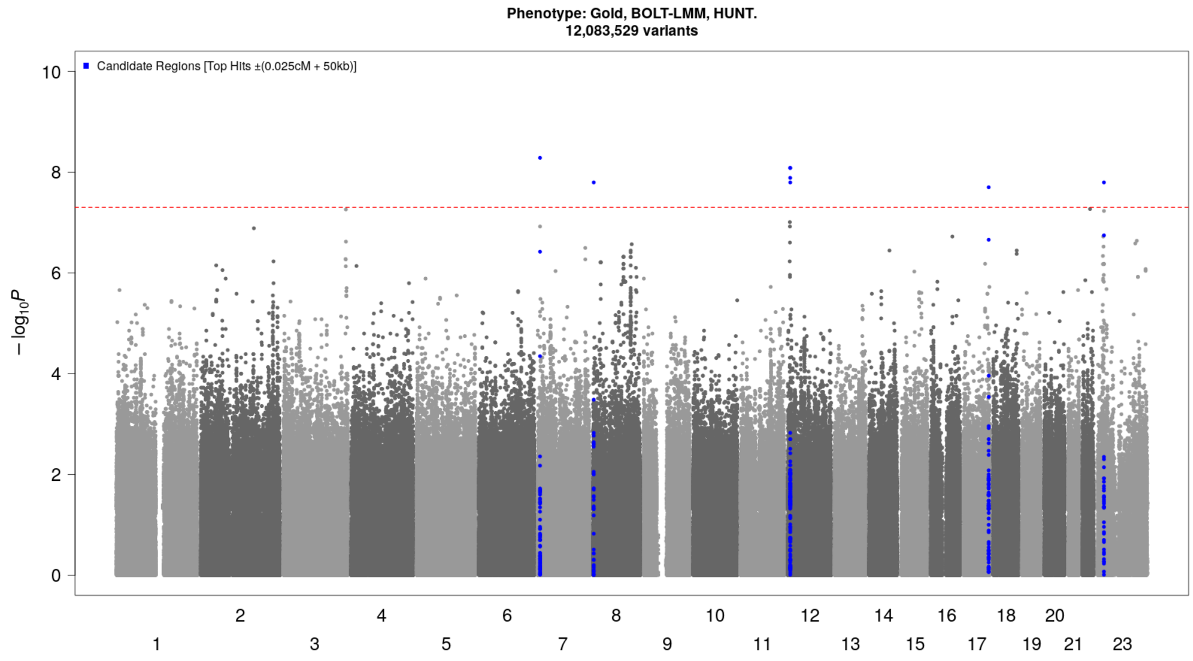


***Supplementary Figure 17: Manhattan plot [gold]:*** *Novel (candidate) genetic loci (index variant ±500 kilobase pairs) associated with whole blood gold concentrations at genome-wide significance (p-value<5×10⁻⁸, threshold indicated by red dotted line) in HUNT. Genetic variants plotted according to chromosome and position (x-axis) and the -log10(p-value) for the variant-gold level association (y-axis). Sample size N=2195. HUNT=The Trøndelag Health Study.*

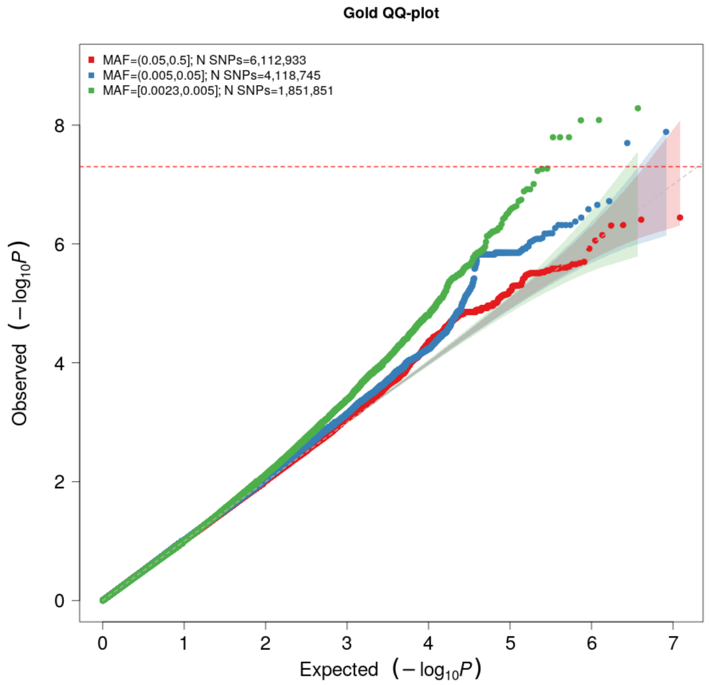


***Supplementary Figure 18: QQ plot [gold].*** *The expected -log10(p-value) given no true associations are given on the x-axis, and the observed -log10(p-value) from the GWA meta-analysis of whole blood gold concentrations in HUNT are given on the y-axis, stratified by minor allele frequency (MAF) categories: Common (MAF>0.05) variants (red), low-frequency (0.005<MAF<0.05) variants (blue) and rare (MAF<0.005) variants (green). HUNT=The Trøndelag Health Study.*

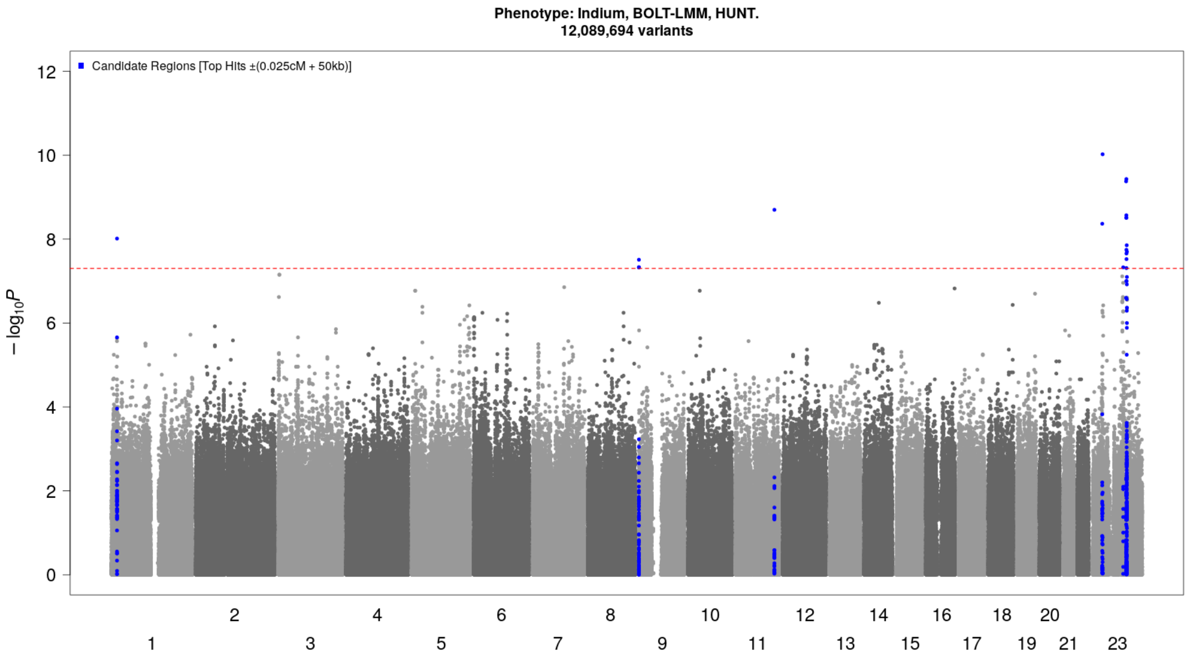


***Supplementary Figure 19: Manhattan plot [indium]:*** *Novel (candidate) genetic loci (index variant ±500 kilobase pairs) associated with whole blood indium concentrations at genome-wide significance (p-value<5×10*$^{-8}$*, threshold indicated by red dotted line) in HUNT. Genetic variants plotted according to chromosome and position (x-axis) and the -log10(p-value) for the variant-indium level association (y-axis). Sample size N=2191. HUNT=The Trøndelag Health Study.*

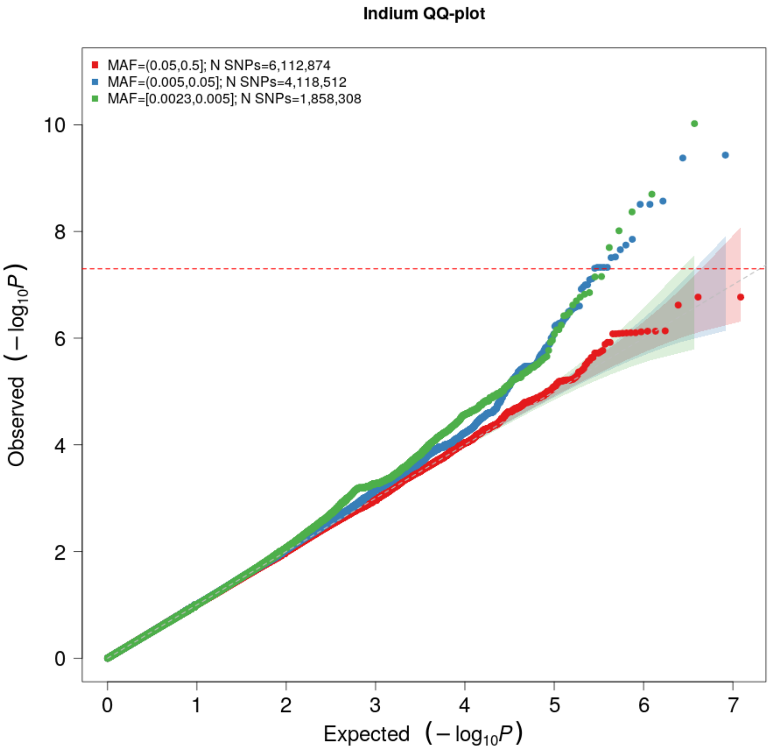


***Supplementary Figure 20: QQ plot [indium].*** *The expected -log10(p-value) given no true associations are given on the x-axis, and the observed -log10(p-value) from the GWA meta-analysis of whole blood indium concentrations in HUNT are given on the y-axis, stratified by minor allele frequency (MAF) categories: Common (MAF>0.05) variants (red), low-frequency (0.005<MAF<0.05) variants (blue) and rare (MAF<0.005) variants (green). HUNT=The Trøndelag Health Study.*

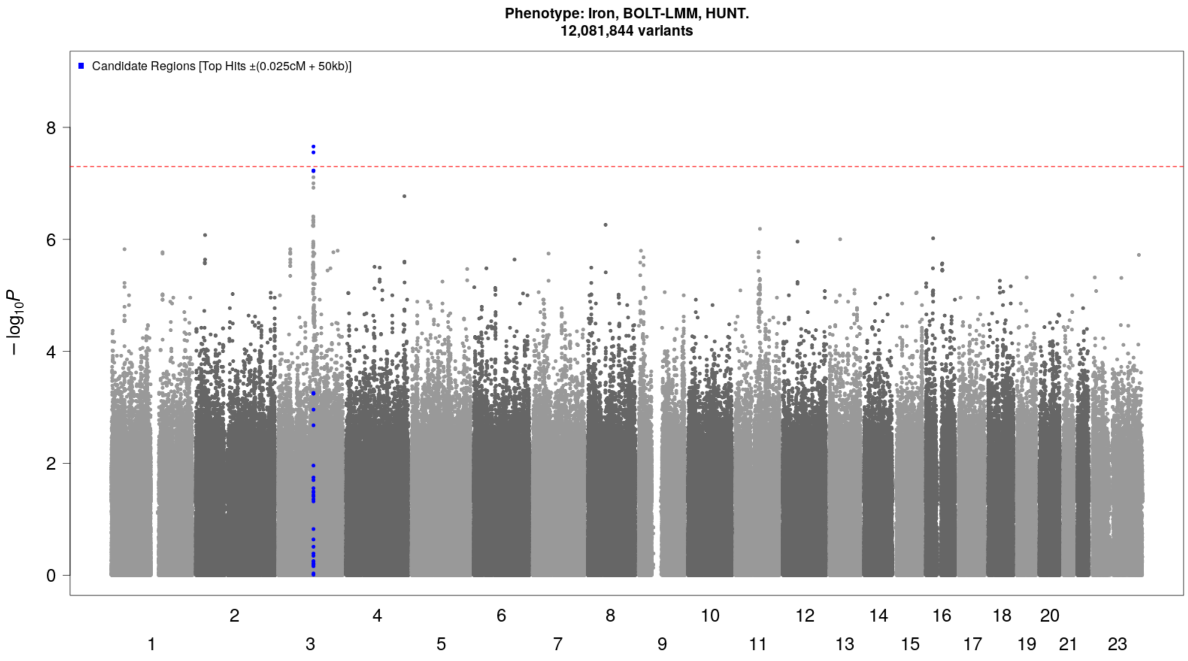


***Supplementary Figure 21: Manhattan plot [iron]:*** *Novel (candidate) genetic loci (index variant ±500 kilobase pairs) associated with whole blood iron concentrations at genome-wide significance (p-value<5×10$^{-8}$, threshold indicated by red dotted line) in HUNT. Genetic variants plotted according to chromosome and position (x-axis) and the -log10(p-value) for the variant-iron level association (y-axis). Sample size N=2197. HUNT=The Trøndelag Health Study.*

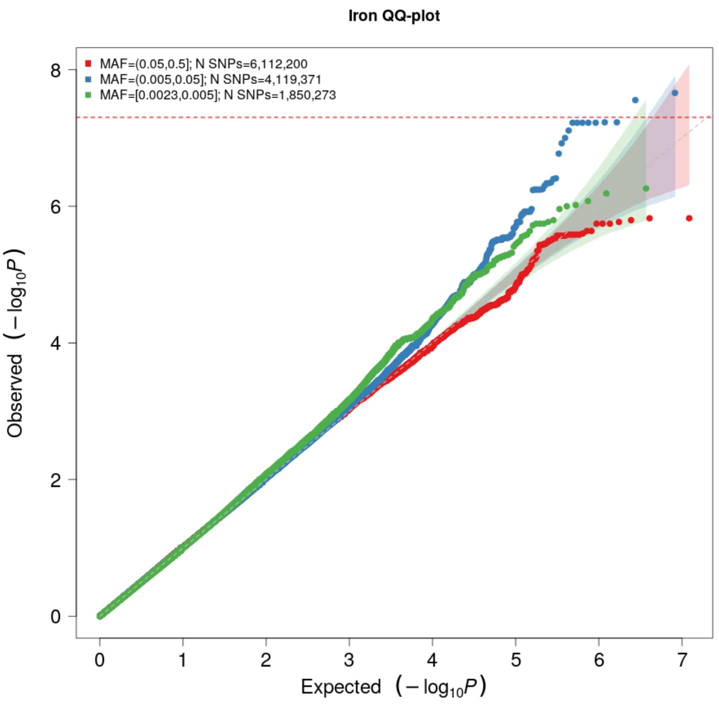


***Supplementary Figure 22: QQ plot [iron]:*** *The expected -log10(p-value) given no true associations are given on the x-axis, and the observed -log10(p-value) from the GWAS of iron concentrations in HUNT are given on the y-axis, stratified by minor allele frequency (MAF) categories: Common (MAF>0.05) variants (red), low-frequency (0.005<MAF<0.05) variants (blue) and rare (MAF<0.005) variants (green). HUNT=The Trøndelag Health Study.*

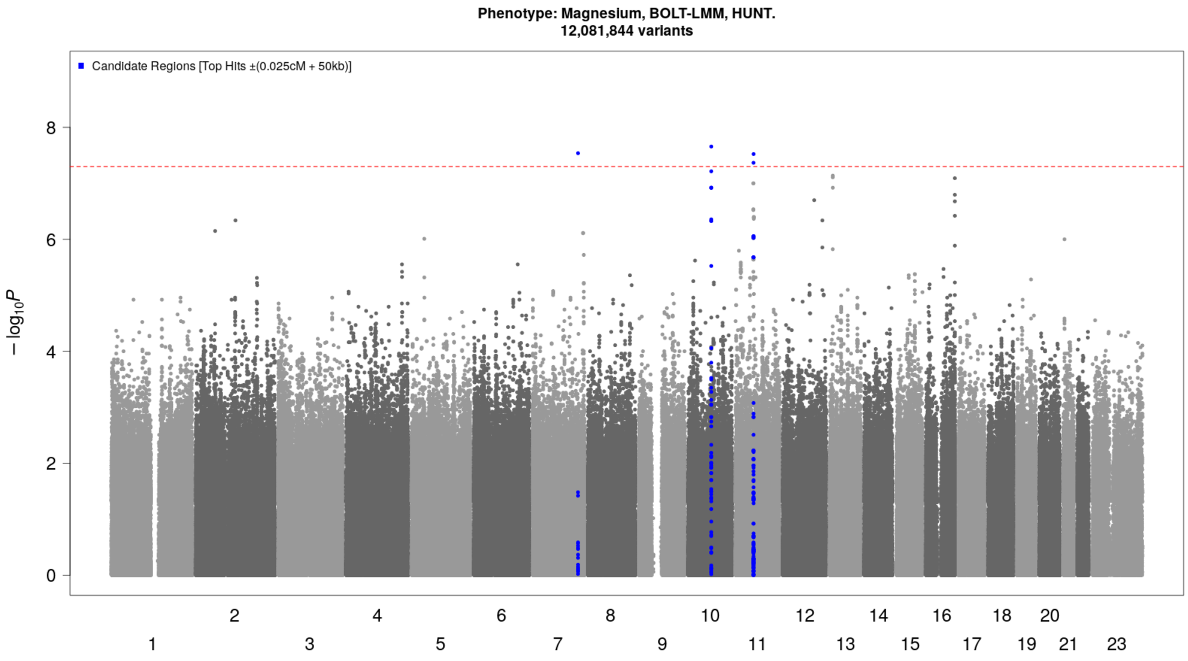


***Supplementary Figure 23: Manhattan plot [magnesium]:*** *Novel (candidate) genetic loci (index variant ±500 kilobase pairs) associated with whole blood magnesium concentrations at genome-wide significance (p-value<5×10⁻⁸, threshold indicated by red dotted line) in HUNT. Genetic variants plotted according to chromosome and position (x-axis) and the -log10(p-value) for the variant-magnesium level association (y-axis). Sample size N=2197. HUNT=The Trøndelag Health Study.*

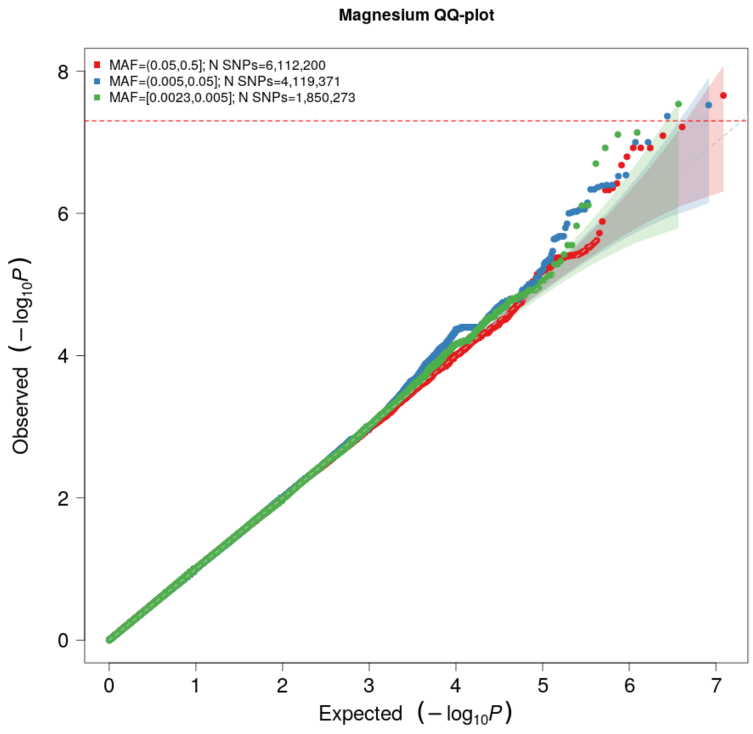


***Supplementary Figure 24: QQ plot [magnesium]:*** *The expected -log10(p-value) given no true associations are given on the x-axis, and the observed -log10(p-value) from the GWAS of magnesium concentrations in HUNT are given on the y-axis, stratified by minor allele frequency (MAF) categories: Common (MAF>0.05) variants (red), low-frequency (0.005<MAF<0.05) variants (blue) and rare (MAF<0.005) variants (green). HUNT=The Trøndelag Health Study.*

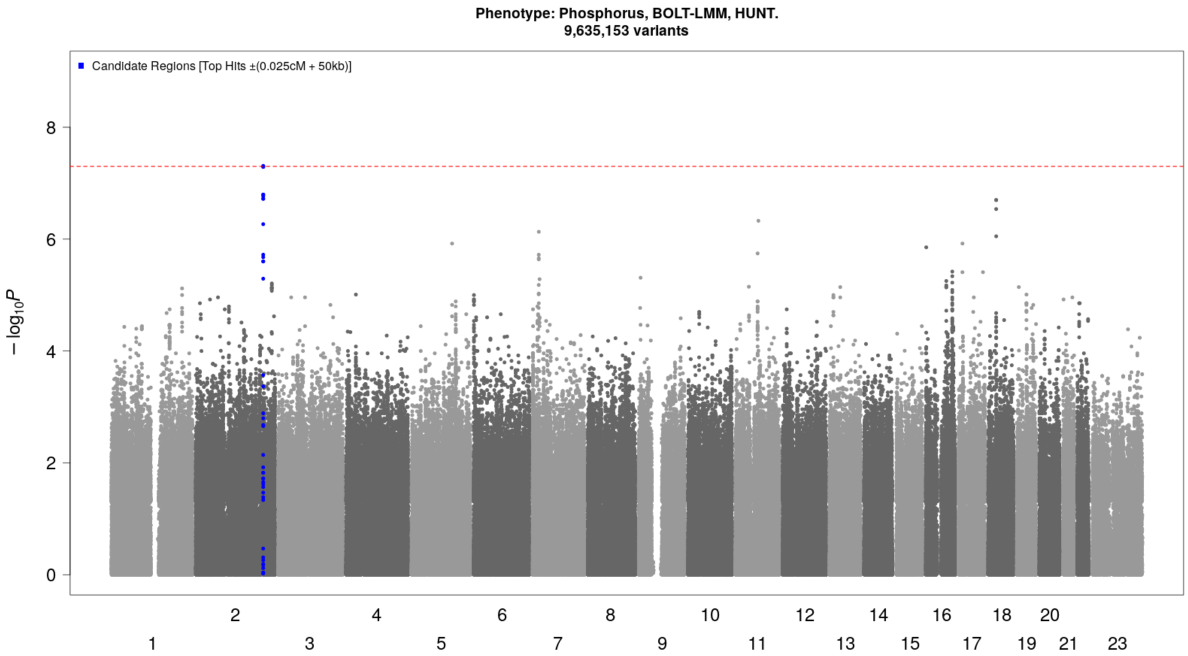


***Supplementary Figure 25: Manhattan plot [phosphorus]:*** *Novel (candidate) genetic loci (index variant ±500 kilobase pairs) associated with whole blood phosphorus concentrations at genome-wide significance (p-value<5×10⁻⁸, threshold indicated by red dotted line) in HUNT. Genetic variants plotted according to chromosome and position (x-axis) and the -log10(p-value) for the variant-phosphorus level association (y-axis). Sample size N=748. HUNT=The Trøndelag Health Study.*

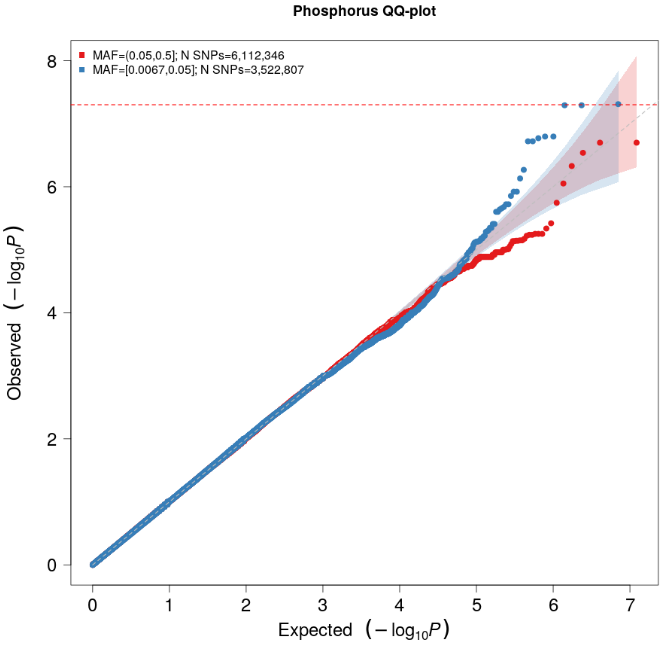


***Supplementary Figure 26: QQ plot [phosphorus]:*** *The expected -log10(p-value) given no true associations are given on the x-axis, and the observed -log10(p-value) from the GWAS of phosphorus concentrations in HUNT are given on the y-axis, stratified by minor allele frequency (MAF) categories: Common (MAF>0.05) variants (red) and low-frequency (0.005<MAF<0.05) variants (blue). HUNT=The Trøndelag Health Study.*

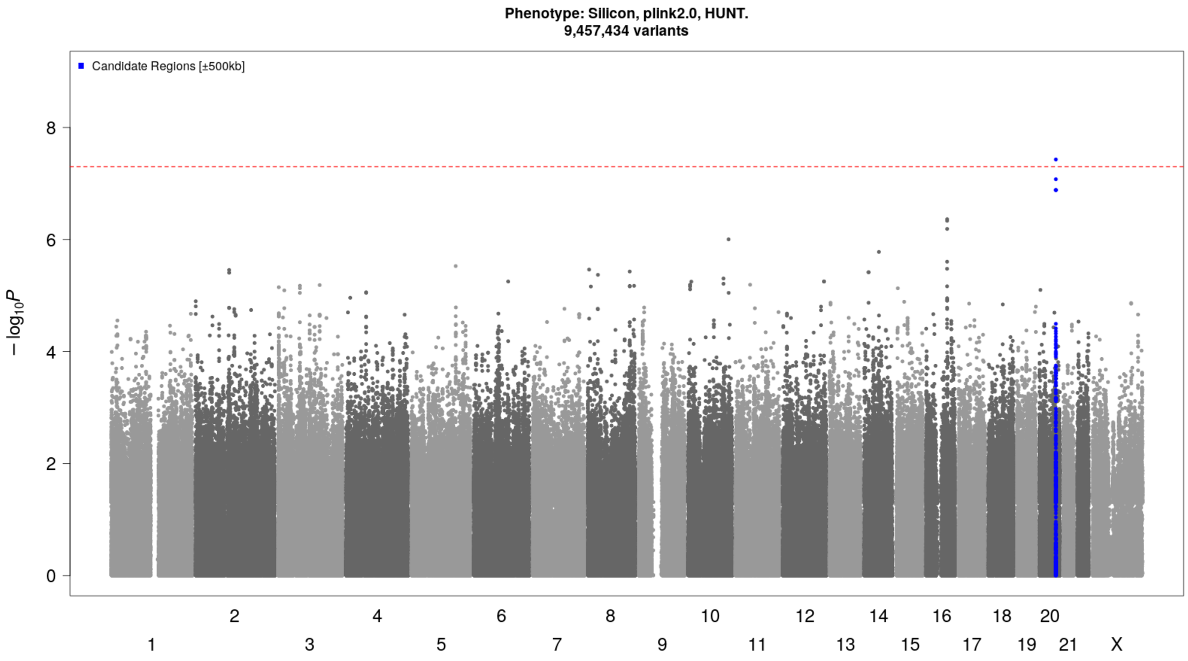


***Supplementary Figure 27: Manhattan plot [silicon]:*** *Novel (candidate) genetic loci (index variant ±500 kilobase pairs) associated with whole blood silicon concentrations at genome-wide significance (p-value<5×10⁻⁸, threshold indicated by red dotted line) in HUNT. Genetic variants plotted according to chromosome and position (x-axis) and the -log10(p-value) for the variant-silicon level association (y-axis). Sample size N=688. HUNT=The Trøndelag Health Study.*

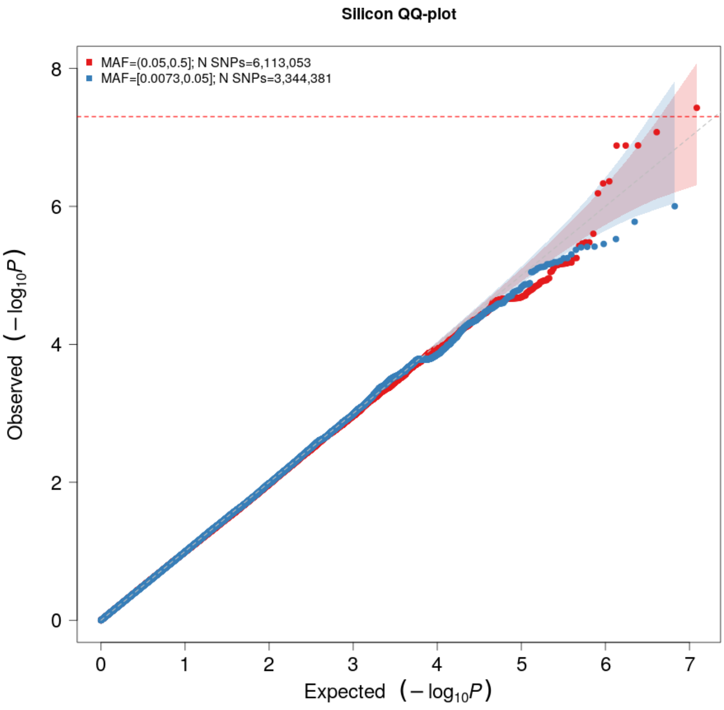


***Supplementary Figure 28: QQ plot [silicon]:*** *The expected -log10(p-value) given no true associations are given on the x-axis, and the observed -log10(p-value) from the GWAS of silicon concentrations in HUNT are given on the y-axis, stratified by minor allele frequency (MAF) categories: Common (MAF>0.05) variants (red) and low-frequency (0.005<MAF<0.05) variants (blue). HUNT=The Trøndelag Health Study.*

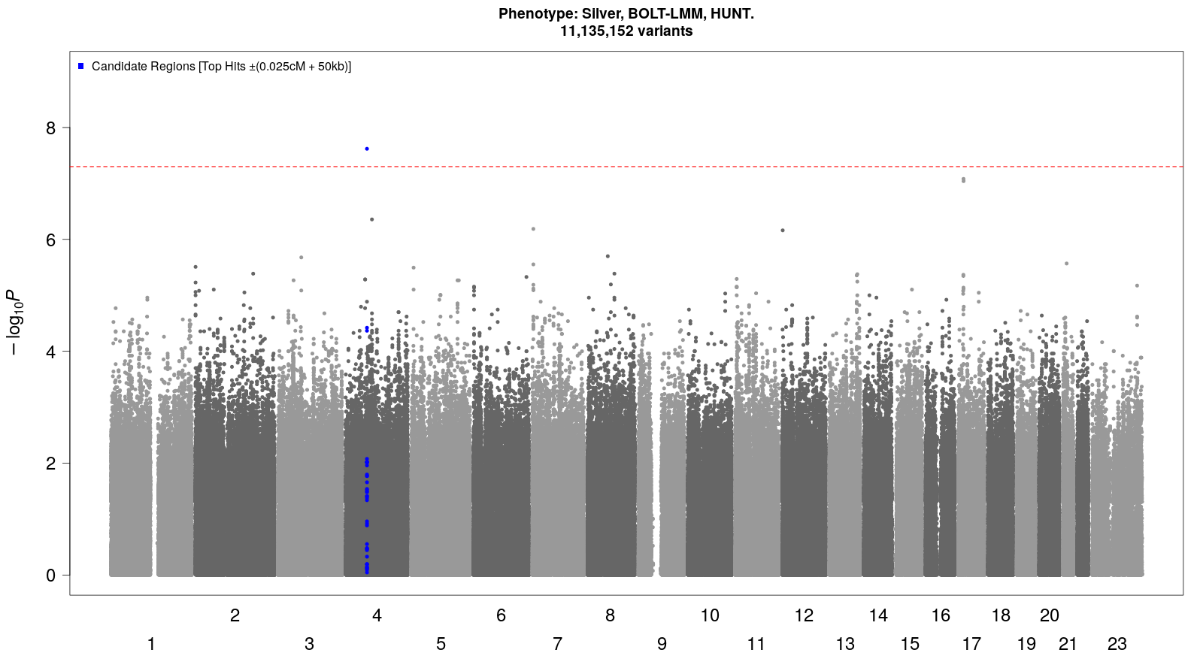


***Supplementary Figure 29: Manhattan plot [silver]:*** *Novel (candidate) genetic loci (index variant ±500 kilobase pairs) associated with whole blood silver concentrations at genome-wide significance (p-value<5×10⁻⁸, threshold indicated by red dotted line) in HUNT. Genetic variants plotted according to chromosome and position (x-axis) and the -log10(p-value) for the variant-silver level association (y-axis). Sample size N=1479. HUNT=The Trøndelag Health Study.*

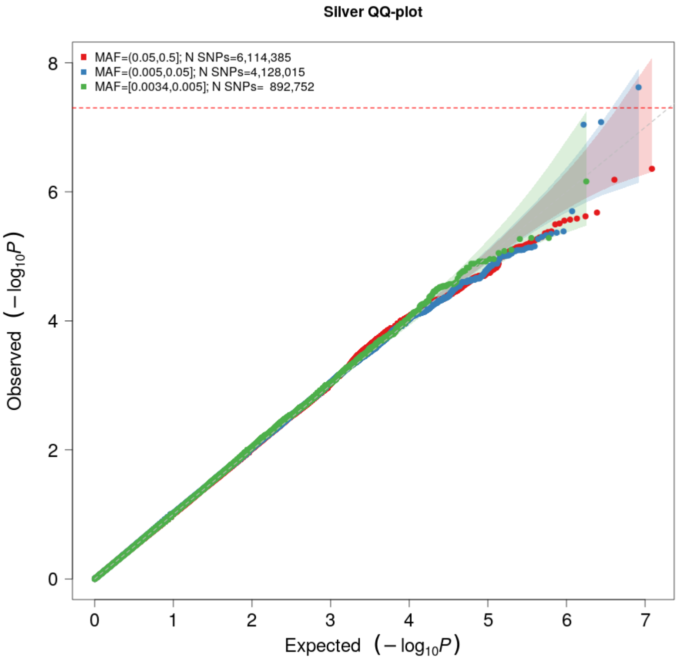


***Supplementary Figure 30: QQ plot [silver]:*** *The expected -log10(p-value) given no true associations are given on the x-axis, and the observed -log10(p-value) from the GWAS of silver concentrations in HUNT are given on the y-axis, stratified by minor allele frequency (MAF) categories: Common (MAF>0.05) variants (red), low-frequency (0.005<MAF<0.05) variants (blue) and rare (MAF<0.005) variants (green). HUNT=The Trøndelag Health Study.*

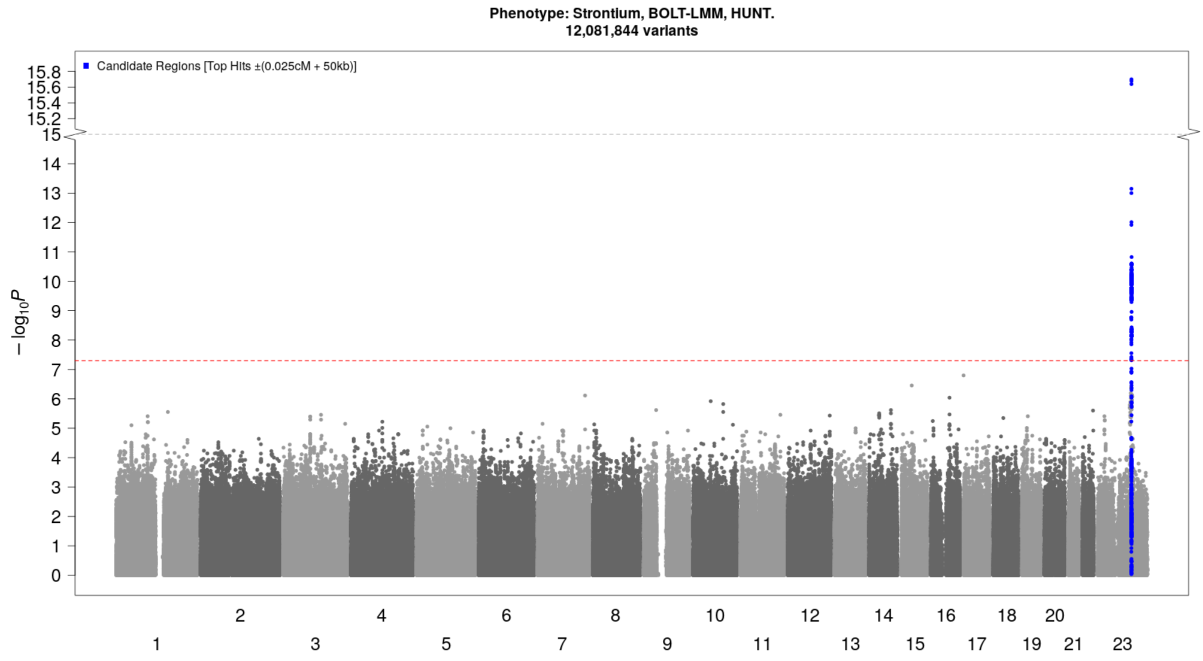


***Supplementary Figure 31: Manhattan plot [strontium]:*** *Novel (candidate) genetic loci (index variant ±500 kilobase pairs) associated with whole blood strontium concentrations at genome-wide significance (p-value<5×10$^{-8}$, threshold indicated by red dotted line) in HUNT. Genetic variants plotted according to chromosome and position (x-axis) and the -log10(p-value) for the variant-strontium level association (y-axis). Sample size N=2197. HUNT=The Trøndelag Health Study.*

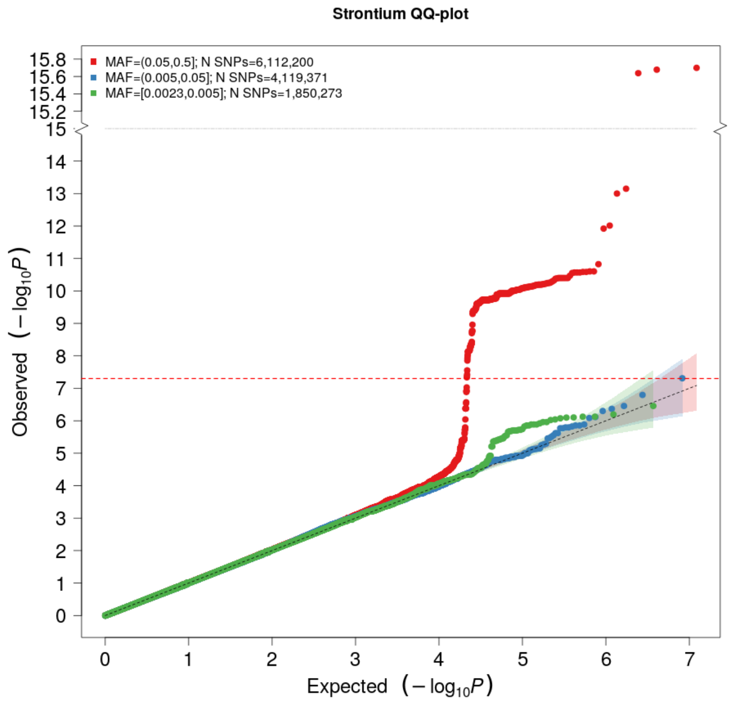


***Supplementary Figure 32: QQ plot [strontium]:*** *The expected -log10(p-value) given no true associations are given on the x-axis, and the observed -log10(p-value) from the GWAS of strontium concentrations in HUNT are given on the y-axis, stratified by minor allele frequency (MAF) categories: Common (MAF>0.05) variants (red), low-frequency (0.005<MAF<0.05) variants (blue) and rare (MAF<0.005) variants (green). HUNT=The Trøndelag Health Study.*

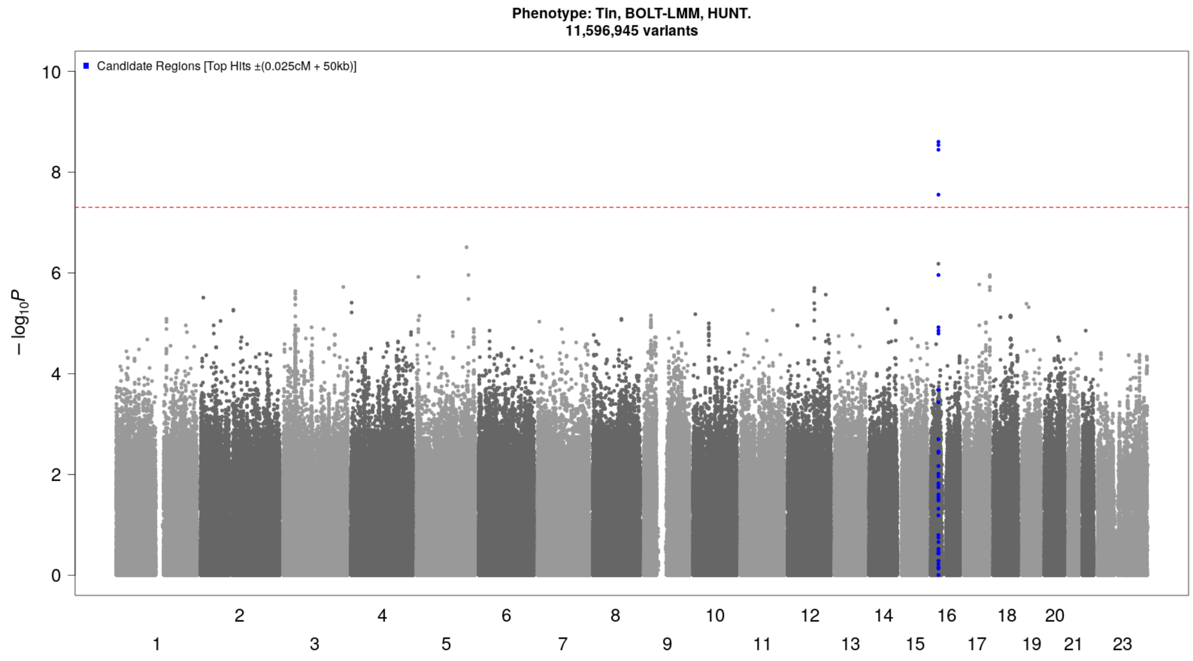


***Supplementary Figure 33: Manhattan plot [tin]:*** *Novel (candidate) genetic loci (index variant ±500 kilobase pairs) associated with whole blood tin concentrations at genome-wide significance (p-value<5×10⁻⁸, threshold indicated by red dotted line) in HUNT. Genetic variants plotted according to chromosome and position (x-axis) and the -log10(p-value) for the variant-tin level association (y-axis). Sample size N=1778. HUNT=The Trøndelag Health Study.*

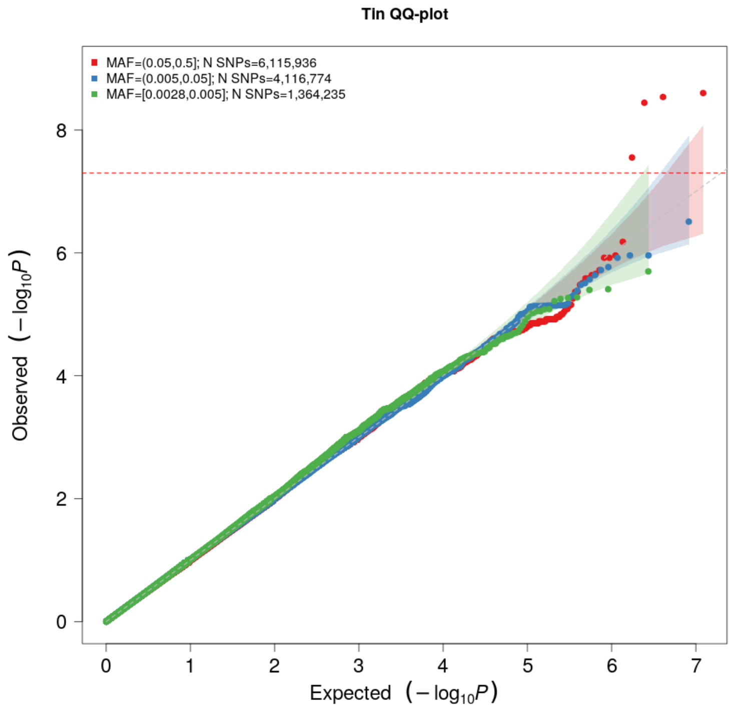


***Supplementary Figure 34: QQ plot [tin]:*** *The expected -log10(p-value) given no true associations are given on the x-axis, and the observed -log10(p-value) from the GWAS of tin concentrations in HUNT are given on the y-axis, stratified by minor allele frequency (MAF) categories: Common (MAF>0.05) variants (red), low-frequency (0.005<MAF<0.05) variants (blue) and rare (MAF<0.005) variants (green). HUNT=The Trøndelag Health Study.*

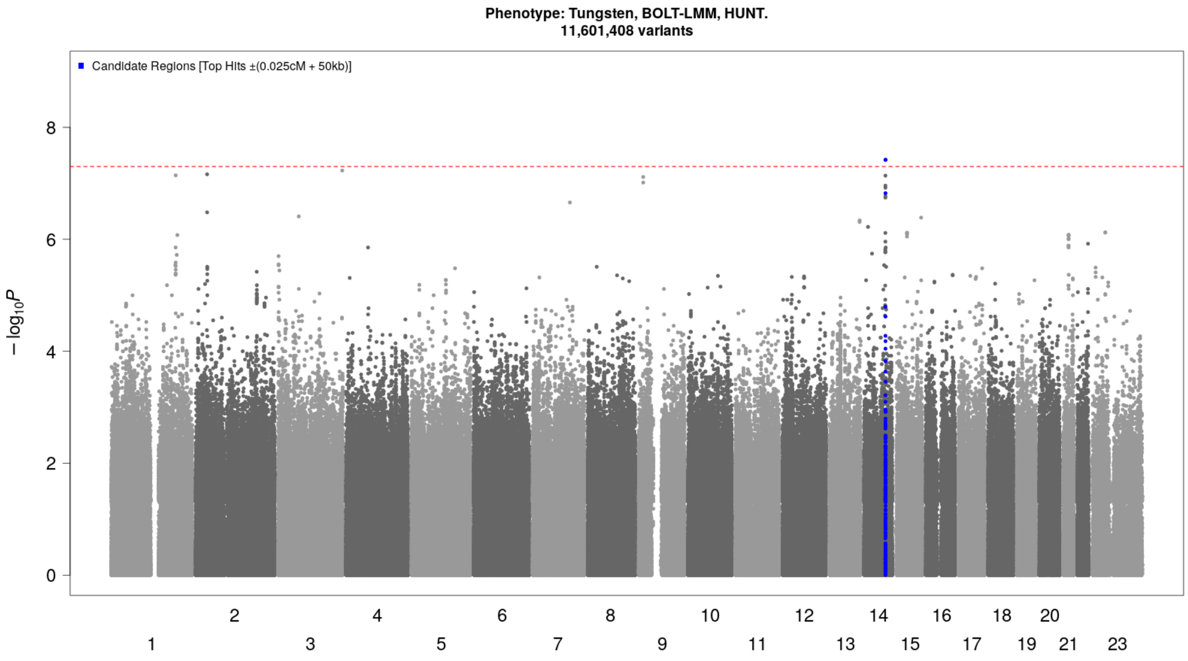


***Supplementary Figure 35: Manhattan plot [tungsten]:*** *Novel (candidate) genetic loci (index variant ±500 kilobase pairs) associated with whole blood tungsten concentrations at genome-wide significance (p-value<5×10⁻⁸, threshold indicated by red dotted line) in HUNT. Genetic variants plotted according to chromosome and position (x-axis) and the -log10(p-value) for the variant-tungsten level association (y-axis). Sample size N=1783. HUNT=The Trøndelag Health Study*

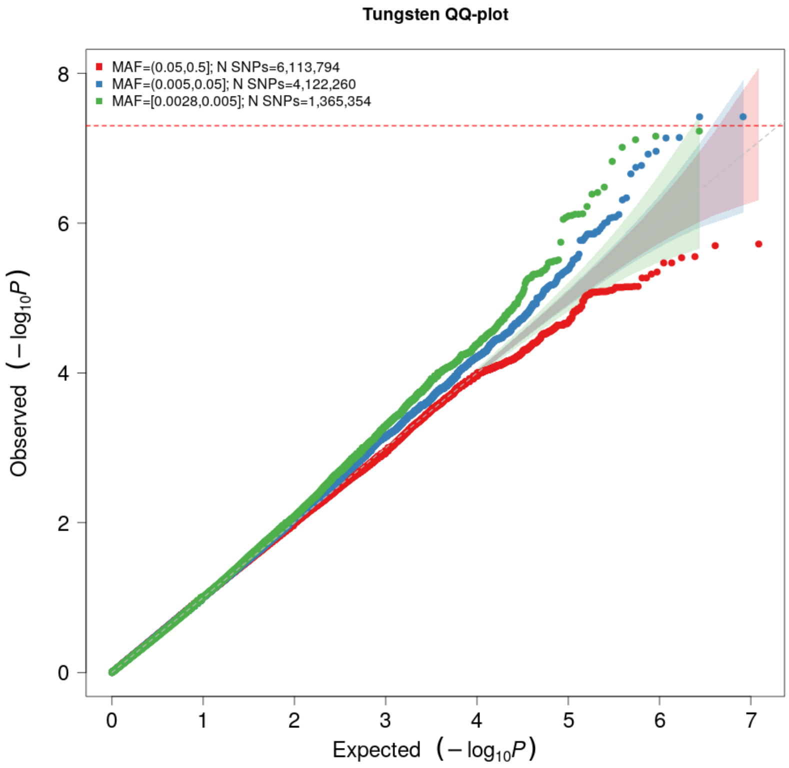


***Supplementary Figure 36: QQ plot [tungsten]:*** *The expected -log10(p-value) given no true associations are given on the x-axis, and the observed -log10(p-value) from the GWAS of tungsten concentrations in HUNT are given on the y-axis, stratified by minor allele frequency (MAF) categories: Common (MAF>0.05) variants (red), low-frequency (0.005<MAF<0.05) variants (blue) and rare (MAF<0.005) variants (green). HUNT=The Trøndelag Health Study.*

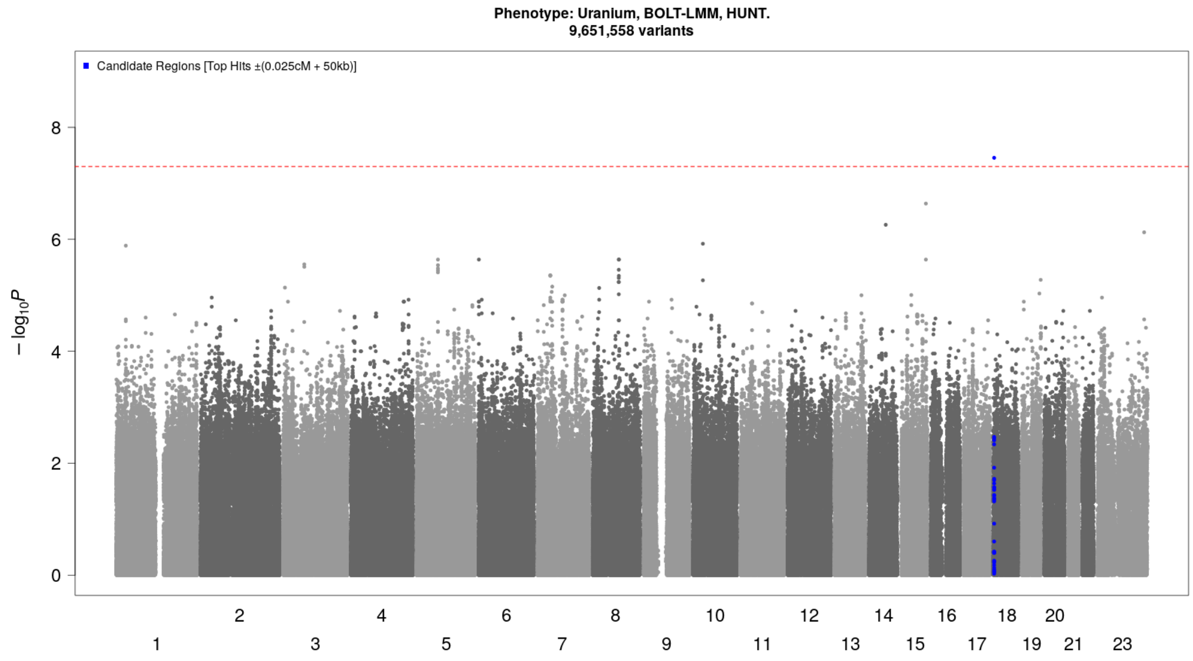


***Supplementary Figure 37: Manhattan plot [uranium]:*** *Novel (candidate) genetic loci (index variant ±500 kilobase pairs) associated with whole blood uranium concentrations at genome-wide significance (p-value<5×10⁻⁸, threshold indicated by red dotted line) in HUNT. Genetic variants plotted according to chromosome and position (x-axis) and the -log10(p-value) for the variant-uranium level association (y-axis). Sample size N=746. HUNT=The Trøndelag Health Study*

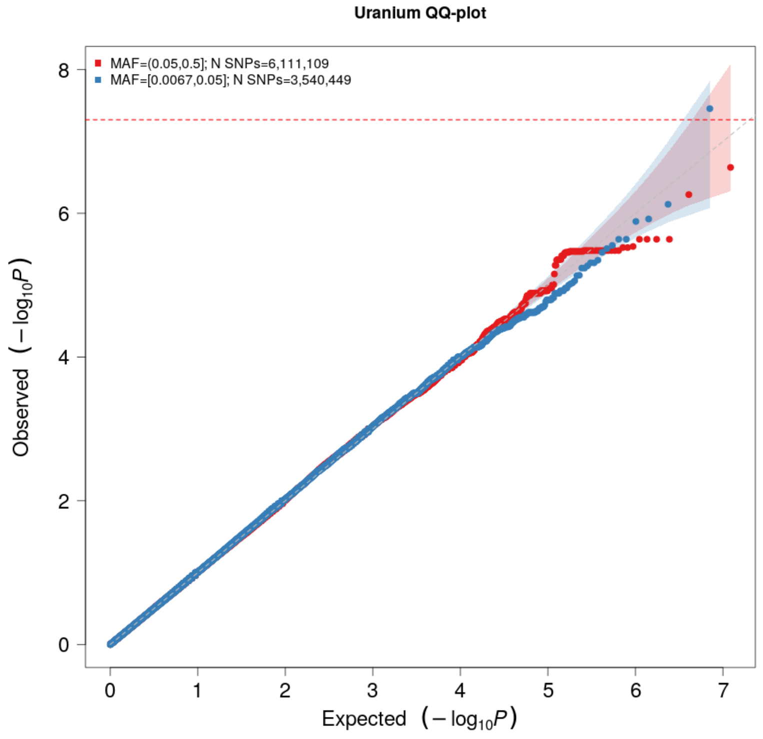


***Supplementary Figure 38: QQ plot [uranium]:*** *The expected -log10(p-value) given no true associations are given on the x-axis, and the observed -log10(p-value) from the GWAS of uranium concentrations in HUNT are given on the y-axis, stratified by minor allele frequency (MAF) categories: Common (MAF>0.05) variants (red) and low-frequency (0.005<MAF<0.05) variants (blue). HUNT=The Trøndelag Health Study.*

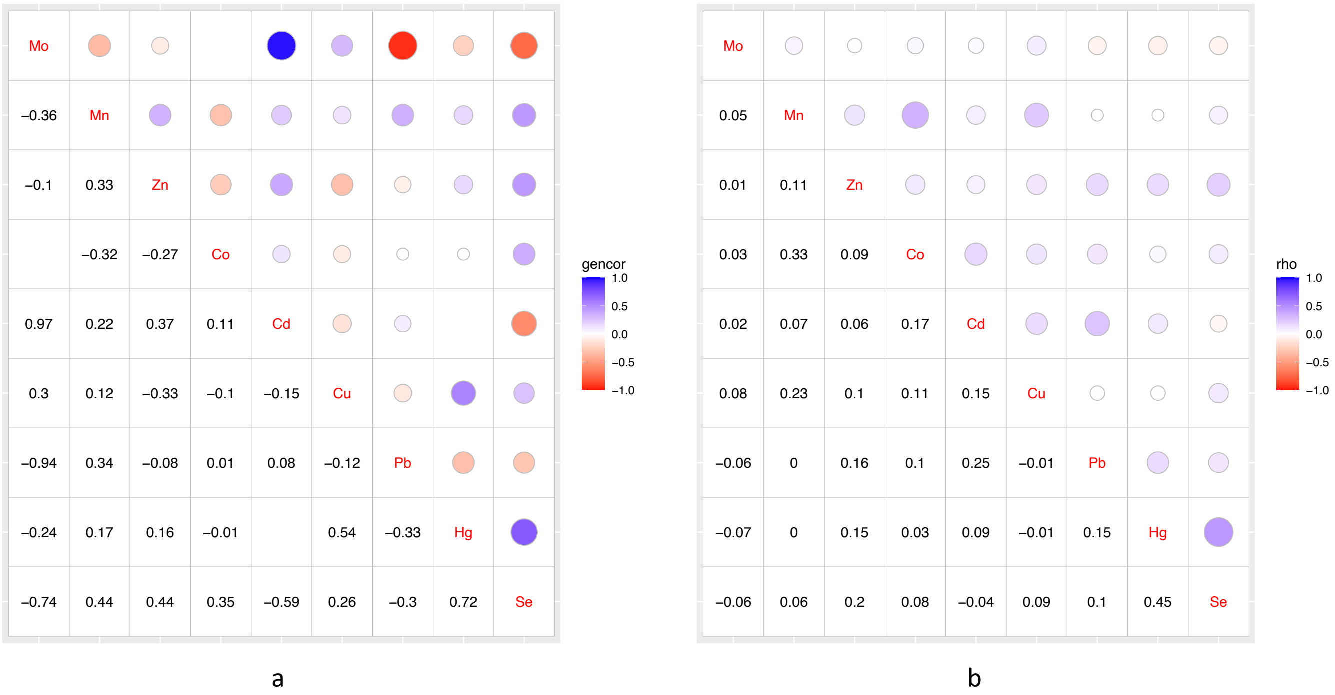


***Supplementary Figure 39: Pairwise correlations between trace elements.*** *The correlations are given on a scale from -1 to +1, where 0 represents no correlation: (a) Genetic correlations (gencor) between nine trace elements with SNP heritability $h^2>0$ and sample size > 5000 estimated by LD Score regression. The confidence intervals of the genetic correlations are given in Supplementary Table 7 (below). (b) Phenotypic correlations (rho) between the concentration of the same trace elements in HUNT (log2 transformed and adjusted by lab median concentration of each trace element) given as the Spearman rank correlation coefficient. The circle size in the upper part of each panel indicates the strength of the correlation, and the color indicates the direction of the correlation (blue=positive, red=negative). Trace elements are ordered according to their binding preferences: the further left in each panel, the more class A properties ("oxygen-seeking"), and the further right, the more class B properties ("nitrogen/sulfur-seeking"). Co=cobalt, Cd=cadmium, Mo=molybdenum, Cu=copper, Hg=mercury, Se=selenium, Zn=zinc, Pb=lead, and Mn=manganese.*

### *Supplementary Table 7: Pairwise phenotypic and genetic correlations of meta-analyzed trace elements*

| Trace element 1 | Trace element 2 | rg | SE | N_rg1 | N_rg2 | Z | P-value | rs | N_rs |
|---|---|---|---|---|---|---|---|---|---|
| Cobalt | Copper | -0.10 | 0.31 | 5052 | 6580 | -0.31 | 0.76 | 0.11 | 1291 |
| Cobalt | Lead | 0.01 | 0.40 | 5052 | 6580 | 0.03 | 0.97 | 0.10 | 1291 |
| Cobalt | Manganese | -0.32 | 0.27 | 5052 | 6580 | -1.20 | 0.23 | 0.33 | 1291 |
| Cobalt | Mercury | -0.01 | 2.53 | 5052 | 6579 | 0.00 | 1.00 | -0.01 | 2819 |
| Cobalt | Selenium | 0.35 | 0.47 | 5052 | 5631 | 0.75 | 0.45 | 0.08 | 1291 |
| Cobalt | Zinc | -0.27 | 0.29 | 5052 | 6580 | -0.95 | 0.34 | 0.09 | 1291 |
| Copper | Lead | -0.12 | 0.33 | 6580 | 6580 | -0.36 | 0.72 | -0.01 | 2819 |
| Copper | Manganese | 0.12 | 0.21 | 6580 | 6580 | 0.57 | 0.57 | 0.23 | 2819 |
| Copper | Mercury | 0.54 | 2.18 | 6580 | 6579 | 0.25 | 0.80 | -0.01 | 2819 |
| Copper | Molybdenum | 0.30 | 0.54 | 6580 | 5746 | 0.55 | 0.58 | 0.08 | 1999 |
| Copper | Selenium | 0.26 | 0.35 | 6580 | 5631 | 0.75 | 0.45 | 0.09 | 2819 |
| Copper | Zinc | -0.33 | 0.31 | 6580 | 6580 | -1.06 | 0.29 | 0.10 | 2819 |
| Cadmium | Cobalt | 0.11 | 0.56 | 6542 | 5052 | 0.20 | 0.84 | 0.17 | 1270 |
| Cadmium | Copper | -0.15 | 0.53 | 6542 | 6580 | -0.28 | 0.78 | 0.15 | 2796 |
| Cadmium | Lead | 0.08 | 0.60 | 6542 | 6580 | 0.14 | 0.89 | 0.25 | 2796 |
| Cadmium | Manganese | 0.22 | 0.36 | 6542 | 6580 | 0.60 | 0.55 | 0.07 | 2796 |
| Cadmium | Molybdenum | 0.97 | 1.80 | 6542 | 5746 | 0.54 | 0.59 | 0.02 | 1982 |
| Cadmium | Selenium | -0.59 | 0.69 | 6542 | 5631 | -0.86 | 0.39 | -0.04 | 2796 |
| Cadmium | Zinc | 0.37 | 0.42 | 6542 | 6580 | 0.89 | 0.38 | 0.06 | 2796 |
| Lead | Manganese | 0.34 | 0.29 | 6580 | 6580 | 1.16 | 0.24 | 0.00 | 2819 |
| Lead | Mercury | -0.33 | 2.49 | 6580 | 6579 | -0.13 | 0.90 | 0.15 | 2819 |
| Lead | Molybdenum | -0.94 | 0.75 | 6580 | 5746 | -1.26 | 0.21 | -0.06 | 1999 |
| Lead | Selenium | -0.30 | 0.51 | 6580 | 5631 | -0.58 | 0.56 | 0.10 | 2819 |
| Lead | Zinc | -0.08 | 0.30 | 6580 | 6580 | -0.27 | 0.79 | 0.16 | 2819 |
| Manganese | Mercury | 0.17 | 0.94 | 6580 | 6579 | 0.18 | 0.86 | 0.00 | 2819 |
| Manganese | Molybdenum | -0.36 | 0.50 | 6580 | 5746 | -0.73 | 0.47 | 0.05 | 1999 |
| Manganese | Selenium | 0.44 | 0.26 | 6580 | 5631 | 1.72 | 0.09 | 0.06 | 2819 |
| Manganese | Zinc | 0.33 | 0.20 | 6580 | 6580 | 1.63 | 0.10 | 0.11 | 2819 |
| Mercury | Molybdenum | -0.24 | 0.84 | 6579 | 5746 | -0.29 | 0.77 | -0.07 | 1999 |
| Mercury | Selenium | 0.73 | 1.91 | 6579 | 5631 | 0.38 | 0.70 | 0.45 | 2819 |
| Mercury | Zinc | 0.16 | 1.68 | 6579 | 6580 | 0.10 | 0.92 | 0.15 | 2819 |
| Molybdenum | Selenium | -0.74 | 0.90 | 5746 | 5631 | -0.82 | 0.41 | -0.06 | 1999 |
| Molybdenum | Zinc | -0.10 | 0.50 | 5746 | 6580 | -0.20 | 0.84 | 0.01 | 1999 |
| Selenium | Zinc | 0.44 | 0.28 | 5631 | 6580 | 1.55 | 0.12 | 0.20 | 2819 |

Genetic and phenotypic correlations for pairs of trace elements with a positive SNP heritability estimate and meta-analysis sample size > 5000. Genetic correlations (rg) with standard error (SE) are estimated by LD Score regression and given with the corresponding z score (Z) and p-value. The genetic correlations for trace elements 1 and 2 are estimated from summary statistics with N_rg1 and N_rg2 individuals, respectively. The phenotypic correlations are estimated from N_rs individuals in HUNT. Phenotypic correlations are estimated as the Spearman rank correlation (rs) of the log2 transformed trace element concentrations corrected for the corresponding lab's median measurement.

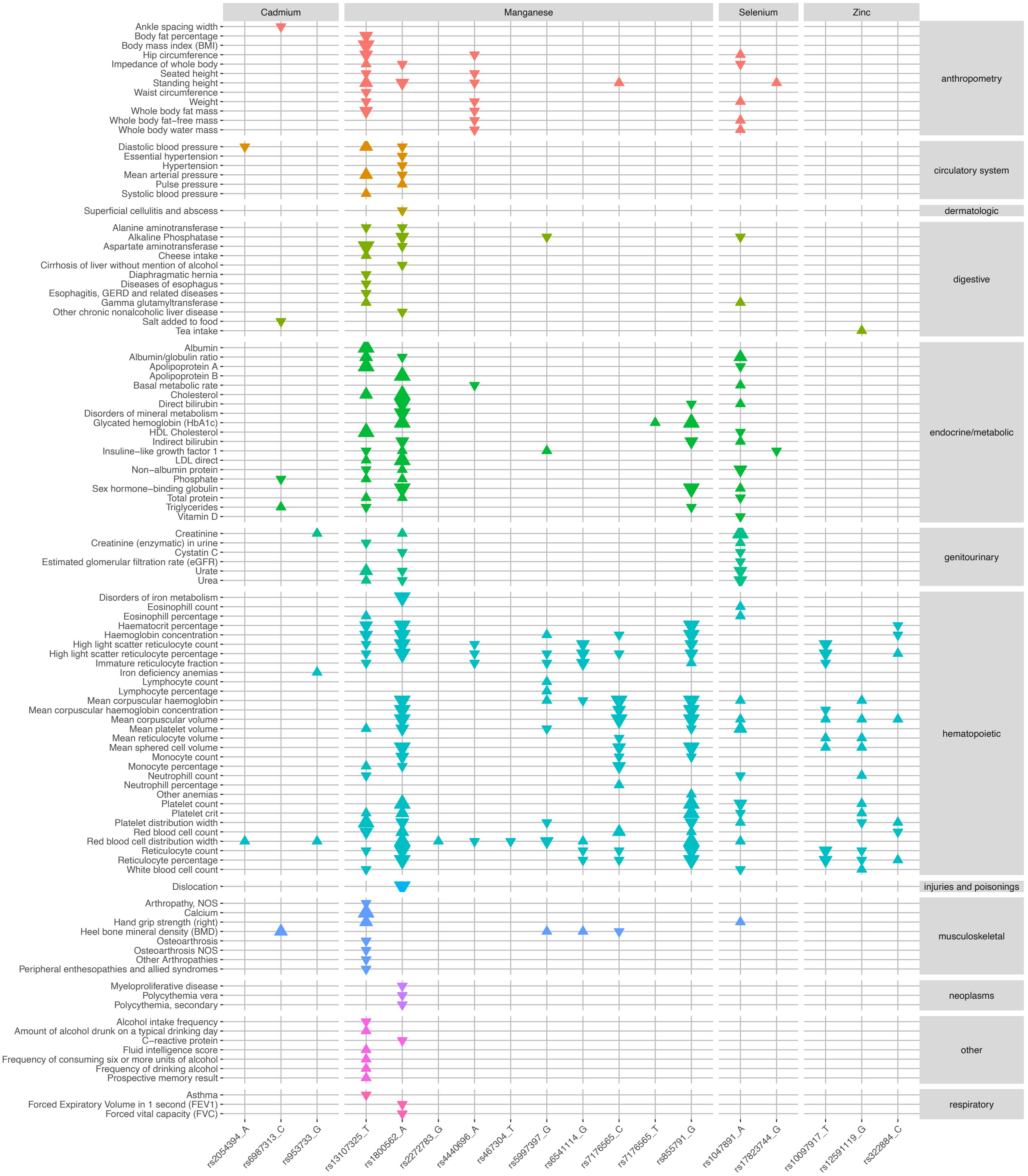


***Supplementary Figure 40: Phenome-wide associations between meta-analysis index variants and phecodes, continuous traits and biomarkers in the UK Biobank.*** *Each triangle represents a statistically significant (p-value<9.7×10$^{-7}$) association between an index variant from trace element GWA meta-analysis (x-axis, grouped by trace element) and an outcome in the UK Biobank (y-axis, grouped and colored according to biological domain). Larger triangles represent lower p-values, and the direction indicates if the direction of effect for the indicated allele is the same (up) or opposite (down) as the association with the trace element.*

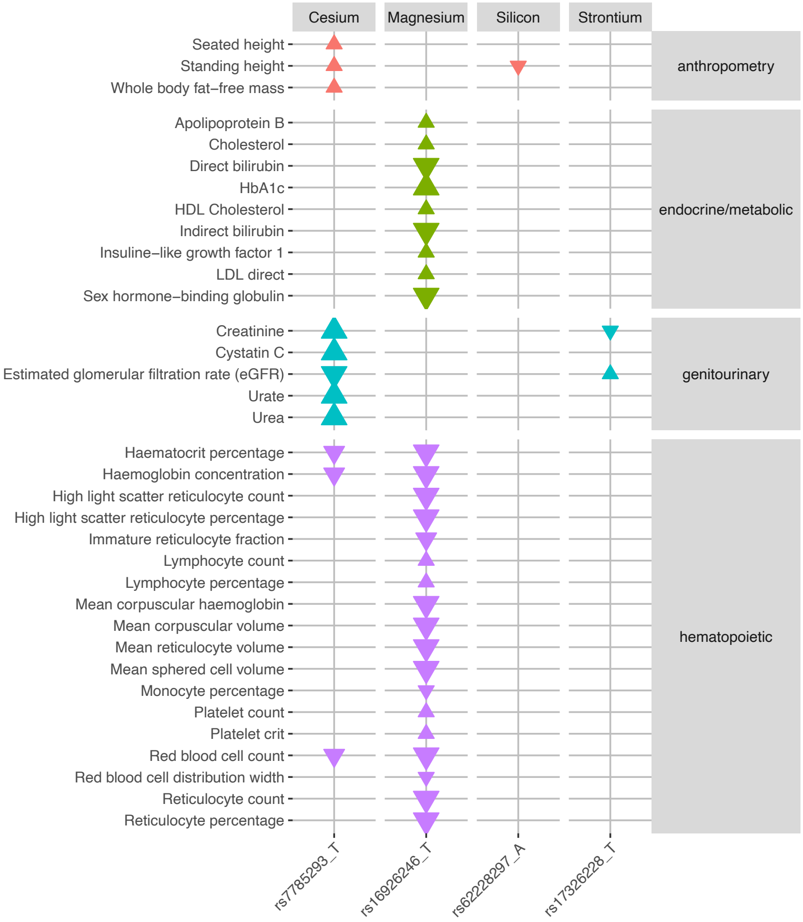


***Supplementary Figure 41: Phenome-wide associations between index variants from GWAS in HUNT and phecodes, continuous traits and biomarkers in the UK Biobank.*** *Each triangle represents a statistically significant (p-value<$9.7 \times 10^{-7}$) association between an index variant from trace element GWASs in HUNT (x-axis, grouped by trace element) and an outcome in the UK Biobank (y-axis, grouped and colored according to biological domain). Larger triangles represent lower p-values, and the direction indicates if the direction of effect for the indicated allele is the same (up) or opposite (down) as the association with the trace element.*

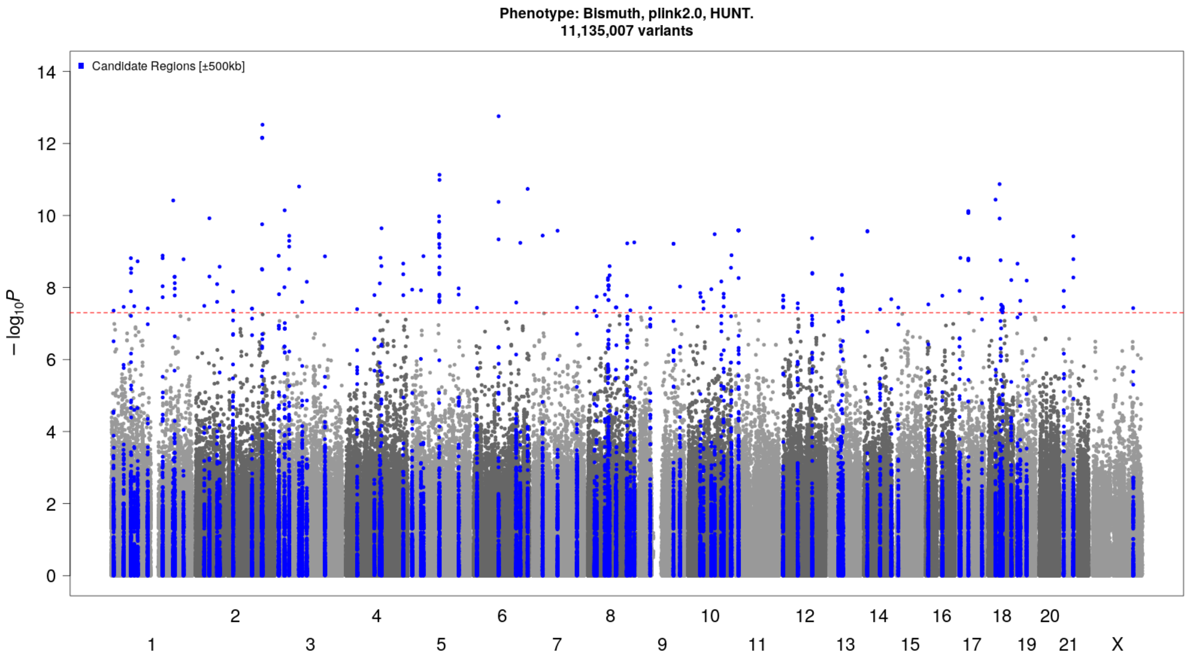


***Supplementary Figure 42: Manhattan plot [bismuth (excluded)]:*** *Inflated test statistics of bismuth concentrations give a high number of apparently associated (index variant ±500 kilobase pairs) genetic loci at genome-wide significance (p-value<5×10⁻⁸, threshold indicated by red dotted line) in HUNT, and the results were therefore excluded. Genetic variants plotted according to chromosome and position (x-axis) and the -log10(p-value) for the variant-bismuth level association (y-axis). Sample size N=1539. HUNT=The Trøndelag Health Study.*

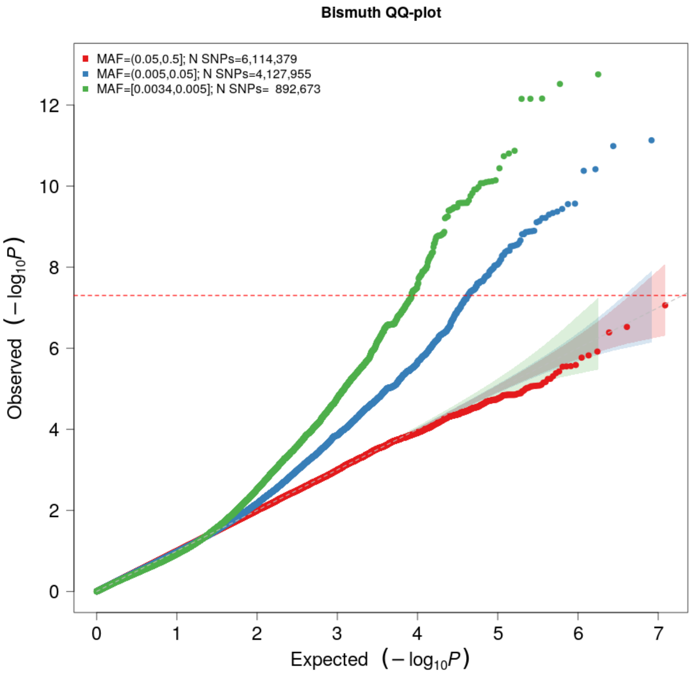


***Supplementary Figure 43: QQ plot [bismuth (excluded)]:*** *The expected -log10(p-value) given no true associations are given on the x-axis, and the observed -log10(p-value) from the GWAS of bismuth concentrations in HUNT are given on the y-axis, stratified by minor allele frequency (MAF) categories: Common (MAF>0.05) variants (red), low-frequency (0.005<MAF<0.05) variants (blue) and rare (MAF<0.005) variants (green). Deviations from the expected values at small -log10(p-values) indicate highly inflated test statistics for low-frequency and rare variants. HUNT=The Trøndelag Health Study.*

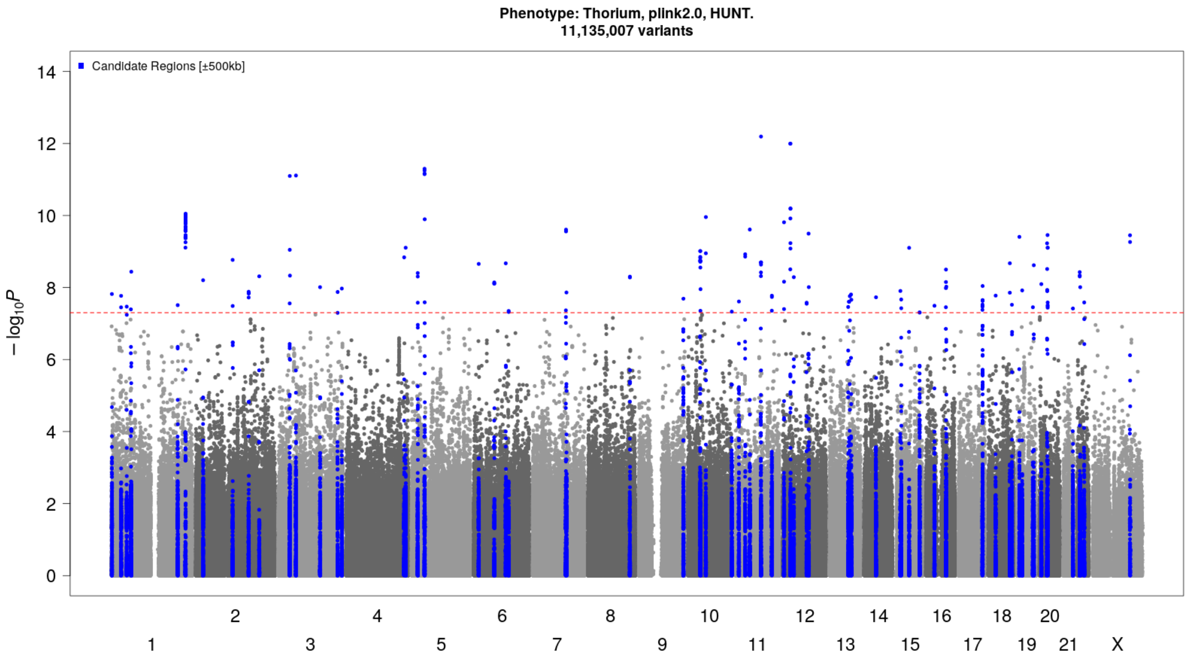


***Supplementary Figure 44: Manhattan plot [thorium (excluded)]:*** *Inflated test statistics of thorium concentrations give a high number of apparently associated (index variant ±500 kilobase pairs) genetic loci at genome-wide significance (p-value<5×10⁻⁸, threshold indicated by red dotted line) in HUNT, and the results were therefore excluded. Genetic variants plotted according to chromosome and position (x-axis) and the -log10(p-value) for the variant-thorium level association (y-axis). Sample size N=1479. HUNT=The Trøndelag Health Study.*

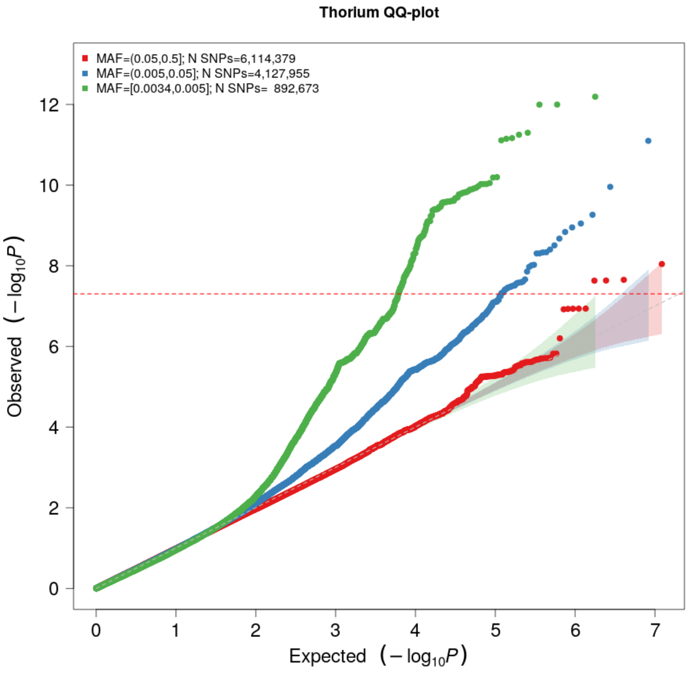


***Supplementary Figure 45: QQ plot [thorium (excluded)]:*** *The expected -log10(p-value) given no true associations are given on the x-axis, and the observed -log10(p-value) from the GWAS of thorium concentrations in HUNT are given on the y-axis, stratified by minor allele frequency (MAF) categories: Common (MAF>0.05) variants (red), low-frequency (0.005<MAF<0.05) variants (blue) and rare (MAF<0.005) variants (green). Deviations from the expected values at small -log10(p-values) indicate highly inflated test statistics for low-frequency and rare variants. HUNT=The Trøndelag Health Study.*
